# Longitudinal analysis of the utility of liver biochemistry in hospitalised COVID-19 patients as prognostic markers

**DOI:** 10.1101/2020.09.15.20194985

**Authors:** Tingyan Wang, David A Smith, Cori Campbell, Steve Harris, Hizni Salih, Kinga A Várnai, Kerrie Woods, Theresa Noble, Oliver Freeman, Zuzana Moysova, Thomas Marjot, Gwilym J Webb, Jim Davies, Eleanor Barnes, Philippa C Matthews

## Abstract

**Background:** COVID-19, the clinical syndrome caused by infection with SARS-CoV-2, has been associated with deranged liver biochemistry in studies from China, Italy and the USA. However, the clinical utility of liver biochemistry as a prognostic marker of outcome for COVID-19 is currently debated.

**Methods:** We extracted routinely collected clinical data from a large teaching hospital in the UK, matching 585 hospitalised SARS-CoV-2 RT-PCR-positive patients to 1165 hospitalised SARS-CoV-2 RT-PCR-negative patients for age, gender, ethnicity and pre-existing comorbidities. Liver biochemistry was compared between groups over time to determine whether derangement was associated with outcome.

**Results:** 26.8% (157/585) of COVID-19 patients died, compared to 11.9% (139/1165) in the non-COVID-19 group (p<0.001). At presentation, a significantly higher proportion of the COVID-19 group had elevated alanine aminotransferase (20.7% vs. 14.6%, p=0.004) and hypoalbuminaemia (58.7% vs. 35.0%, p<0.001), compared to the non-COVID-19 group. Within the COVID-19 group, those with hypoalbuminaemia at presentation had 1.83-fold increased hazards of death compared to those with normal albumin (adjusted hazard ratio [HR] 1.83, 95% CI 1.25-2.67), whilst the hazard of death was ~4-fold higher in those aged ≥75 years (adjusted HR 3.96, 95% CI 2.59-6.04) and ~3-fold higher in those with pre-existing liver disease (adjusted HR 3.37, 95% CI 1.58-7.16). In the COVID-19 group, alkaline phosphatase increased (R=0.192, p<0.0001) and albumin declined (R=-0.123, p=0.0004) over time in patients who died. We did not find a significant association between other liver biochemistry and death.

**Conclusion:** In this UK population, liver biochemistry is commonly deranged in patients with COVID-19 but only baseline low albumin and a rising alkaline phosphatase over time are prognostic markers for death.

**Key Points:**

- Age ≥ 75 years and low albumin at the time of a positive SARS-CoV-2 RT-PCR test can predict poor clinical outcome in COVID-19 patients.
- Liver biochemistry is more likely to be abnormal in patients with COVID-19 than in patients without COVID-19.
- Patients with COVID-19 who died showed a greater decline in albumin and a greater increase in alkaline phosphatase over time, compared to those who survived.
- Patients with pre-existing liver disease and COVID-19 had an increased mortality.

**Lay Summary:** We used routinely collected hospital data from a large UK teaching hospital to compare liver biochemistry (markers of liver inflammation or damage) between 585 patients with COVID-19 and 1165 patients of the same age and sex admitted to hospital but without COVID-19. Patients with COVID-19 were more likely to die than those without COVID-19, and deaths were significantly higher in those aged ≥75 years. We found that patients with COVID-19 were more likely to have abnormal liver biochemistry. Low albumin (a blood protein) at the time of being diagnosed with COVID-19 was associated with an increased chance of death.

## Introduction

Over 28 million confirmed cases of Severe Acute Respiratory Syndrome Coronavirus 2 (SARS-CoV-2) infection and 922,252 deaths have been reported globally as of 14^th^ September 2020; and the UK is one of the worst affected countries with 368,508 confirmed cases and 41,628 deaths reported (1). The clinical syndrome caused by SARS-CoV-2 is COVID-19, which primarily affects the respiratory system, but other organs, including the heart, gastrointestinal tract and liver may be affected, and a systemic sepsis syndrome may develop (2).

Existing data on liver biochemistry in COVID-19 patients have been reported from China, the USA and Italy.. These studies report that 37-69% of patients with COVID-19 had at least one abnormal liver biochemistry on hospital admission (3-6) while 93% had at least one abnormal liver biochemistry over the course of disease (6). Specially, the prevalence estimates of elevated alanine aminotransferase (ALT), aspartate aminotransferase (AST) and bilirubin (BR) in COVID-19 patients are 9%-28%, 14%-35% and 6%-23%, respectively (3-9). Some studies reported that liver biochemistry abnormalities are associated with longer hospital stay (4), or clinical severity (3, 10), whereas other studies have not found a relationship between liver biochemistry and severity (7, 9). The set of liver biochemistry reported for COVID-19 patients varies: ALT, AST, and total BR are typically included with alkaline phosphatase (ALP) and, Gamma-glutamyl transferase (GGT) less frequently reported.

Albumin is a non-specific marker of liver function, and has been less consistently assessed; it is typically reported as one parameter of patients’ baseline characteristics, with limited investigation of its impact as a prognostic marker. A recent meta-analysis of 20 retrospective studies from China revealed that patients with severe COVID-19 had lower albumin level compared to mild cases, but there was significant heterogeneity between studies (11). Another meta-analysis showed that hypoalbuminaemia could be included in prognostic machine learning models to predict severe COVID-19 or mortality (12).

Several studies have investigated potential associations between liver biochemistry and death in COVID-19 patients (6-8), or included liver biochemistry in the development of predictive models (13-16). A report from Italy showed that ALP >150 U/L at admission (without adjusting for relevant confounders) was associated with clinical deterioration in 292 COVID-19 patients (7), and another study from the USA reported that peak ALT >5 times upper limit of normal (ULN) during admission was associated with death in a cohort of 2,273 COVID-19 patients (8). However, another USA study reported that elevations in ALT and AST elevation on admission were associated with length of stay, ICU admission and intubation but not death (6). Studies from Wuhan, China did not find associations of ALT (13, 14) or AST (14) elevation on admission with death in COVID-19 patients, whilst several studies have reported associations of elevated total bilirubin (14) and low albumin on admission (14-16) with risk of death have been reported. Given these variable associations between liver biochemistry and COVID-19 outcomes, the prognostic value of liver biochemistry derangement in COVID-19 needs further evaluation.

Having established a clinical data pipeline through the National Institute for Health Research (NIHR) Health Informatics Collaborative (HIC) (17, 18), our tertiary referral hospital in the UK is strongly placed to undertake analyses using electronic health data from hospitalised patients. Using this resource, we aimed to determine the prevalence of deranged liver biochemistry at baseline and over the disease course in COVID-19 patients, with comparison to a matched group of non-COVID-19 patients admitted during the same period. We also aimed to determine whether baseline liver biochemistry derangement was associated with risk of death in COVID-19 patients, and to compare longitudinal changes in liver biochemistries between COVID-19 patients who died and who survived.

## Methods

### Data collection

We used routinely collected clinical data from Oxford University Hospitals (OUH) NHS Foundation Trust, a large teaching hospital trust in the South East of the UK, with ~1000 inpatient beds. The data is collected by the local NIHR HIC team in Oxford, being drawn automatically from operational systems into a data warehouse and linked to produce a comprehensive record for each patient, as previously described (17). All of the data used for this studied were provided in anonymised form by OUH NHS Foundation Trust, with the prior approval of the Trust Information Governance Team, following the satisfactory completion of a Data Protection Impact Assessment. The management of the dataset is governed by the NIHR HIC Data Sharing Framework. Research Ethics Committee approval was obtained for the database and associated activity: IRAS ID 174658; REC reference 15/SC/0523.

The data extracted for this study included detailed information on demographics, body mass index (BMI), emergency admissions, blood test results, diagnostic codes, procedures, intensive care unit admission, prescriptions, medicines administration, and discharge destination/outcome for all patients admitted to OUH between 1^st^ January 2020 and 21^st^ August 2020.

### Inclusion and exclusion criteria

To select eligible data for adults with/without COVID-19, the inclusion criteria were: (a) at least one RT-PCR nose/throat swab having been undertaken (COVID-19 patients were defined by at least one positive result; COVID-19-negative patients were defined by the absence of a positive result); (b) age ≥ 18 years when tested; (c) hospitalised patients; and (d) at least one episode of liver biochemistry recorded at the time of receiving RT-PCR test or after RT-PCR test (the set of liver biochemistry routinely tested by OUH clinical biochemistry laboratory comprises ALT, ALP, albumin, and BR; AST and GGT are not routinely tested). Exclusion criteria were: (a) SARS-CoV-2 RT-PCR test results reported as invalid; (b) missing age; and (c) pregnancy (antenatal/delivery/post-partum).

### Definitions

We defined baseline as the date of the first positive SARS-CoV-2 RT-PCR test for a COVID-19 patient and the date of the first negative test for a patient without COVID-19. The end of follow-up was defined as the date of last available electronic patient record data for a patient.

The normal ranges set by the hospital biochemistry lab are: ALT: 10-45 IU/L; ALP: 30-130 IU/L; BR: 0-21 umol/L; Albumin: 32-50 g/L. The reference ranges for other blood tests are provided in **Table S1**. We defined baseline liver biochemistry (ALT, ALP, BR, or albumin) as the liver biochemistry measured within 7 days of SARS-CoV-2 RT-PCR test and baseline derangement as at least one abnormal result at this time point. We defined peak/nadir liver biochemistry derangement as at least one abnormal value at any point during follow-up. We defined liver biochemistry recovery as normalisation following derangement. The primary outcome was death during follow-up, and secondary outcomes included ICU admission and invasive ventilation.

Pre-existing comorbidities were defined by a historical diagnosis of a disease before baseline, and diagnosis codes retrieved were provided in **Table S2**. A full list of prescribed drugs searched for data extraction is provided in **Table S3**.

### Statistical analysis

Statistical analysis was performed using R version 4.0.2. All significance tests performed were two-sided. *p* values <0.05 were deemed statistically significant.

#### Propensity Score Matching

We selected eligible subjects, and conducted propensity score matching process to ensure the COVID-19 group was comparable to the non-COVID-19 group in terms of demographics and pre-existing conditions. We used the following variables to calculate propensity scores: age, gender, ethnicity, and pre-existing comorbidities (liver disease, diabetes mellitus (DM), hypertension (HTN), coronary heart disease (CHD), chronic kidney disease (CKD), and cancer). We performed propensity score matching using the package *MatchIt*, with the nearest-neighbour method applied, the matching ratio and calliper size set as 1:2 and 0.1 respectively, and without replacement.

#### Comparison of COVID-19 and matched non-COVID-19 patients

For continuous variables, we calculated median and interquartile range (IQR), or mean and standard deviation (SD), and used Wilcoxon test or t-test for comparison. For categorical variables, we computed number and percentage, and used chi-square or Fisher’s exact test for comparison. We used the Shapiro-Wilk test and graphical methods for normality check.

#### Investigation on whether liver biochemistry predict outcomes in COVID-19 patients

We compared the presence of clinical outcomes in the COVID-19 and non-COVID-19 groups. We also performed Kaplan-Meier (K-M) analysis to compare the survival probability over time in COVID-19 and non-COVID-19 groups. Within the COVID-19 and non-COVID-19 groups, we compared demographics, BMI, comorbidities, baseline and peak/nadir liver biochemistry between those who died vs. survived. We then performed K-M analysis to compare the survival probabilities over time between subgroups with and without deranged baseline liver biochemistry. We used univariate and multivariate Cox proportional-hazards models to investigate whether liver biochemistry predicted death, reporting hazard ratios (HRs) and 95% confidential intervals (CIs). To investigate associations of additional patient characteristics with risk of death, and the robustness of HRs to adjustment for additional confounders, we performed sensitivity/subset analysis whereby associations were investigated in a subset of patients who were not missing data for confounders.

#### Longitudinal analysis of liver biochemistry in COVID-19 patients

We compared liver biochemistry between COVID-19 group and non-COVID-19 group at each time point (to examine difference between groups), as well as investigating liver biochemistry changes over time by comparing liver biochemistry at subsequent time points to their baseline within each group (to examine longitudinal changes). Within the COVID-19 group, we performed longitudinal analysis of liver biochemistry in subgroups of patients who died and survived by fitting linear regression lines with 95% CIs, and reporting Pearson’s correlation coefficients and linear regression significance.

## Results

### Identification, demographics, and outcomes of COVID-19 compared to non-COVID-19 groups

We identified 6311 eligible patients (585 adults with SARS-CoV-2 infection and 5726 without) according to the inclusion/exclusion criteria (**Figure S1)**. Based on our 585 COVID-19-positive patients, we matched a cohort of 1165 COVID-19-negative patients. After matching, there were no significant differences in demographics and pre-existing comorbidities between the two groups (**Table 1**; **Table S4**). Median duration of follow-up was 58 [IQR: 14-104] days in the COVID-19 group and 50 [IQR: 20-78] days in the matched non-COVID-19 group, with no difference in the monitoring duration of liver biochemistry (**Table 1**). The distribution of admitting specialties (**Table S5)** and prescribed antiviral therapy did not differ significantly between COVID-19 and non-COVID-19 groups (**Table S6)**.

**Table 1.** Baseline characteristics and outcomes of patients with and without COVID-19. Data are the median [IQR] or number (%) unless otherwise indicated. For categorical variables, Fisher exact test was performed for comparison on cells with small counts (<5), otherwise Chi-square test was used. For continuous variables, Wilcoxon test was used for comparison due to non-normality.^#^ 194 vs. 344 missing on BMI data; BMI categories were reported based on WHO classification. †93 vs. 191 patients with vs. without COVID-19 did not have baseline data available on all the four liver biochemistries. In detail, 93 vs. 191 missing on ALT; 91 vs. 180 missing on ALP; 93 vs. 191 missed data in bilirubin; 89 vs. 177 missing in albumin. ‡158 vs. 303 patients with vs. without COVID-19 did not have baseline data available on blood clotting tests. ^§^ 162 vs. 324 patients with vs. without COVID-19 did not have baseline data available on vital signs. *ALT, Alanine transaminase; ALP, Alkaline phosphatase; APTT, Activated partial thromboplastin time; BMI, Body mass index; CHD, Coronary heart disease; CKD, Chronic kidney disease; CRP, C-reactive protein; DM, Diabetes mellitus; HTN, Hypertension; INR, International normalised ratio; IQR, Interquartile range; LLN, Lower limit of normal; ULN, Upper limit of normal*..

|  | COVID-19 group<br>(n=585) | non-COVID-19 group<br>(n=1165) | p-value |
| --- | --- | --- | --- |
| Overall follow-up duration<br>(median [IQR]), days | 58 [14,104] | 50 [20,78] | **p=0.002** |
| Duration of liver biochemistry<br>monitoring (median [IQR]), days | 38 [7, 79] | 37 [10, 67] | p=0.51 |
| Gender (male), n(%) | 312 (53.3) | 629 (54.0) | 0.83 |
| Age at test (median [IQR]) | 73 [57, 84] | 73 [58, 83] | 0.76 |
| Ethnicity category, n(%) |  |  | 0.92 |
| Asian | 31 (5.3) | 62 (5.3) |  |
| Black | 20 (3.4) | 35 (3.0) |  |
| Mixed | 10 (1.7) | 22 (1.9) |  |
| White | 415 (70.9) | 804 (69.0) |  |
| Other | 9 (1.5) | 23 (2.0) |  |
| Not stated | 100 (17.1) | 219 (18.8) |  |
| BMI(median [IQR]), kg/m<sup>2</sup> | 26 [23.2, 31.0] | 26 [22.7, 30.5] | 0.47 |
| BMI category<sup>#</sup>, n(%) |  |  | 0.31 |
| <18.5 | 17 (4.3) | 40 (4.9) |  |
| ≥18.5 - <25 | 140 (35.8) | 311 (37.9) |  |
| ≥25 - <30 | 115 (29.4) | 238 (29.0) |  |
| ≥30 - <35 | 77 (19.7) | 122 (14.9) |  |
| ≥35 - <40 | 26 (6.6) | 73 (8.9) |  |
| ≥40 | 16 (4.1) | 37 (4.5) |  |
| **Pre-existing comorbidities, n(%)** |  |  |  |
| Liver disease | 17 (2.9) | 33 (2.8) | 1 |
| DM | 88 (15.0) | 175 (15.0) | 1 |
| HTN | 179 (30.6) | 349 (30.0) | 0.83 |
| CHD | 59 (10.1) | 113 (9.7) | 0.86 |
| CKD | 56 (9.6) | 107 (9.2) | 0.86 |
| Cancer | 41 (7.0) | 88 (7.6) | 0.75 |
| **Outcomes** |  |  |  |
| Death, n(%) | 157 (26.8) | 139 (11.9) | **<0.001** |
| ICU admission, n(%) | 71 (12.1) | 50 (4.3) | **<0.001** |
| Used invasive ventilation in ICU, n(%) | 47 (8.0) | 29 (2.5) | **<0.001** |
| **Baseline liver biochemistry<sup>†</sup>** |  |  |  |
| ALT, >ULN, n(%) | 102 (20.7) | 142 (14.6) | **0.004** |
| ALT (median [IQR]), IU/L | 25 [16, 40] | 19 [13, 32] | **<0.001** |
| ALP, >ULN, n(%) | 98 (19.8) | 213 (21.6) | 0.47 |
| ALP (median [IQR]), IU/L | 85 [66, 114] | 90 [71, 124] | **0.005** |
| Bilirubin, >ULN, n(%) | 29 (5.9) | 127 (13.0) | **<0.001** |
| Bilirubin (median [IQR]), umol/L | 9 [6, 13] | 10 [7, 16] | **<0.001** |
| Albumin, <LLN, n (%) | 291 (58.7) | 346 (35.0) | **<0.001** |
| Albumin (median [IQR]), g/L | 30 [27, 34] | 34 [29, 37] | **<0.001** |
| **Baseline blood clotting tests<sup>‡</sup>, median[IQR]** |  |  |  |
| Prothrombin time, seconds | 10.9 [10.4, 11.4] | 10.9 [10.4, 11.8] | 0.093 |
| APTT, seconds | 24.8 [23.0, 27.7] | 24.4 [22.6, 27.1] | **0.014** |
| INR | 1.0 [1.0, 1.1] | 1.0 [1.0, 1.1] | 0.073 |
| **Baseline renal function tests** |  |  |  |
| Creatinine (median [IQR]), umol/L | 82 [65, 111] | 81 [65, 117] | 0.71 |
| Elevated creatinine, n(%) | 151 (29.9) | 321 (31.3) | 0.63 |
| Urea (median [IQR]), mmol/L | 6.3 [4.3, 9.8] | 6.2 [4.5, 9.9] | 0.64 |
| eGFR (median [IQR]), ml/min/1.73m<sup>2</sup> | 76 [49, >90] | 76 [47, >90] | 0.65 |
| eGFR <90 ml/min/1.73m<sup>2</sup>, n(%) | 369 (73.2) | 746 (72.6) | 0.86 |
| **Other tests at baseline, (median[IQR])** |  |  |  |
| CRP, mg/L | 78 [27, 148] | 20 [4, 93] | **<0.001** |
| Platelets, x10<sup>9</sup>/L | 216 [160, 281] | 245 [195, 311] | **<0.001** |
| Lymphocytes, x10<sup>9</sup>/L | 0.9 [0.6, 1.3] | 1.2 [0.8, 1.8] | **<0.001** |
| **Baseline vital signs<sup>§</sup>,**<br>**(median [IQR])** |  |  |  |
| Temperature tympanic, °C | 37 [36.5, 38] | 36.5 [36, 37] | **<0.001** |
| Heart rate, bpm | 86 [76, 97] | 81 [72, 95] | **<0.001** |
| Oxygen saturation, % | 95 [94, 97] | 96 [95, 98] | **<0.001** |
| Respiratory rate | 20 [18, 22] | 18 [17, 19] | **<0.001** |
| Diastolic blood pressure, mmHg | 70 [64, 78] | 72 [64, 81] | 0.076 |
| Systolic blood pressure, mmHg | 129 [118, 143] | 133 [121, 149] | **<0.001** |

Over a median follow-up of 58 days vs. 50 days, 26.8% (157/585) of COVID-19 patients died compared to 11.9% (139/1165) in the non-COVID-19 group (p<0.001), respectively. The COVID-19 group had a higher rate of ICU admission during hospitalisation (12.1% vs. 4.3%, p<0.001), and a higher rate of invasive ventilation use in ICU (8% vs. 2.5%, p<0.001) (**Table 1**). The K-M estimated probability of surviving beyond 30 days after SARS-CoV-2 RT-PCR test was 77% for COVID-19 patients compared to 92% for non-COVID-19 patients (**Figure 1**).

**Figure 1.**
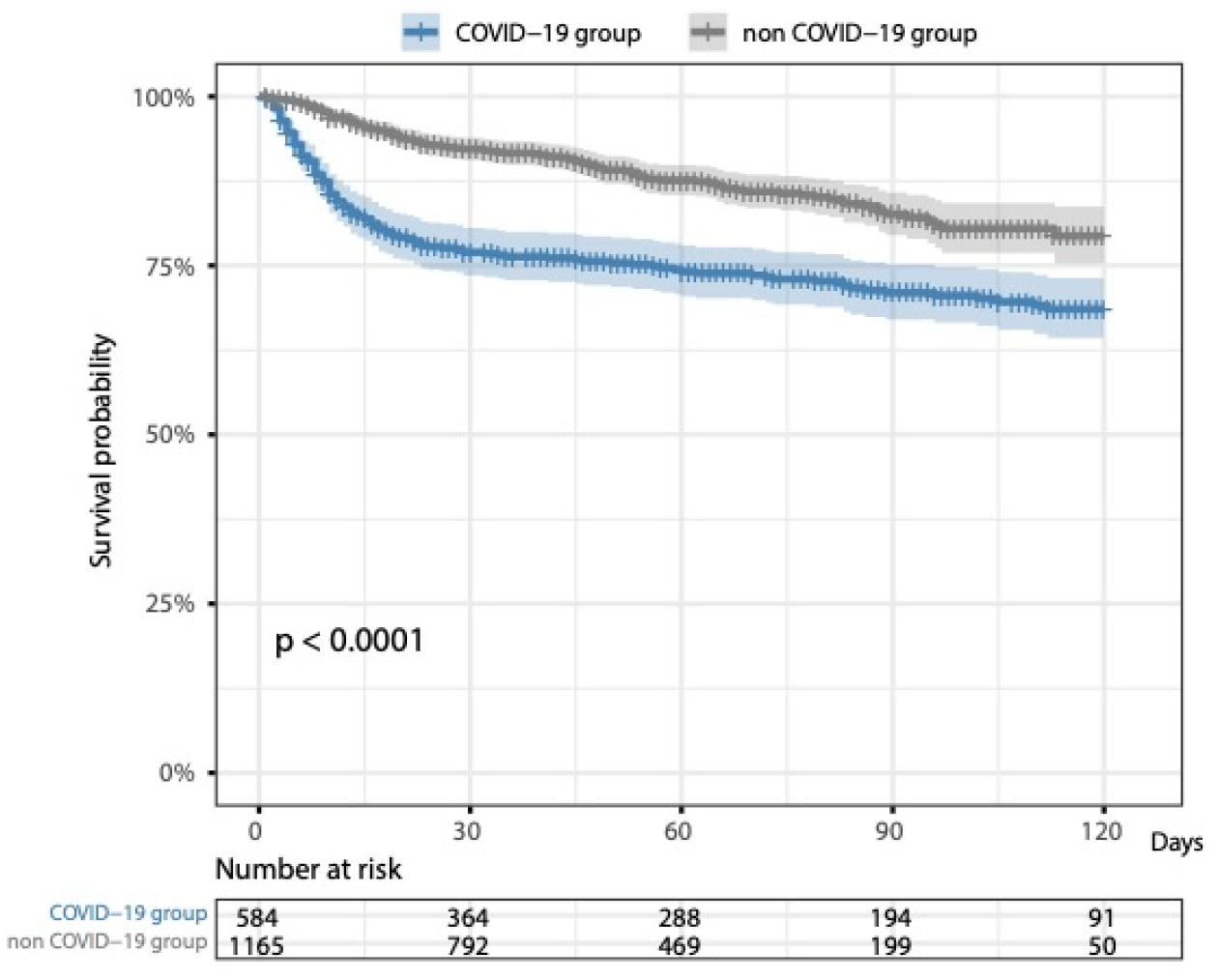
K-M curves for the comparison of time to death in COVID-19 group vs. non-COVID-19 group since SARS-CoV-2 RT-PCR test. *One in COVID-19 group has missing death date. p-value was based on the logrank test*.

### Assessment of liver biochemistry in the COVID-19 compared to non-COVID-19 group

Baseline liver biochemistry was available in 492 COVID-19 patients and 974 non-COVID-19 patients; patients with deranged peak liver biochemistry were less likely to have missing baseline liver biochemistry data (**Table S7)**. The median time interval between consecutive liver biochemistry measurements was 1 [IQR: 1-4] day and 2 [IQR: 1-7] days, for COVID-19 and non-COVID-19 groups respectively.

At baseline, the COVID-19 group had a significantly higher median ALT value (25 IU/L vs. 19 IU/L, p<0.001) and a higher proportion of patients with ALT >ULN (20.7% vs.14.6%, p=0.004) than the non-COVID-19 group. The COVID-19 group also had a lower median albumin (30 g/L vs. 34 g/L, p<0.001), lower platelets (216 x10*9/L vs. 245 x10*9/L, p<0.001), lower lymphocytes (0.9 x10*9/L vs. 1.2 x10*9/L, p<0.001) and a significantly higher CRP (78 mg/L vs. 20 mg/L, p<0.001), as compared to the non-COVID-19 group. Activated partial thromboplastin time (APPT) and baseline vital signs were also more deranged in the COVID-19 group, whilst renal function was preserved (**Table 1**). Overall, the COVID-19 group had a significantly higher proportion of patients with ≥1 deranged liver biochemistry at baseline compared to the non-COVID-19 group (72.6% vs. 55.5%, p<0.001) (**Table S8**).

Over follow-up, the COVID-19 group also had more deranged liver biochemistry, with a higher median peak ALT (34 IU/L vs. 26 IU/L, p<0.001), a higher proportion with peak ALT >ULN (37.9% vs. 27.7%, p<0.001), a lower median nadir albumin (26 g/L vs. 29 g/L, p<0.001), and a higher prevalence of hypoalbuminaemia (79.0% vs. 59.5%, p<0.001) compared to the non-COVID-19 group (**Table S8**). COVID-19 patients also had significantly higher median ALT and lower median albumin values at all time points throughout follow-up (7, 14, 21, and 28 days), compared to the non-COVID-19 group (all p<0.05) (**Figure 2A-B**). In the COVID-19 group, median ALT increased at 7 and 14 days, compared to baseline (both p<0.05) (**Figure 2C**), and median albumin decreased at 7 days of follow-up compared to baseline (p<0.0001) and maintained at low levels at subsequent time points (**Figure 2D**). We did not identify differences in ALP and bilirubin over time between these groups, other than at baseline or at 7 days (**Figure S2)**.

**Figure 2.**
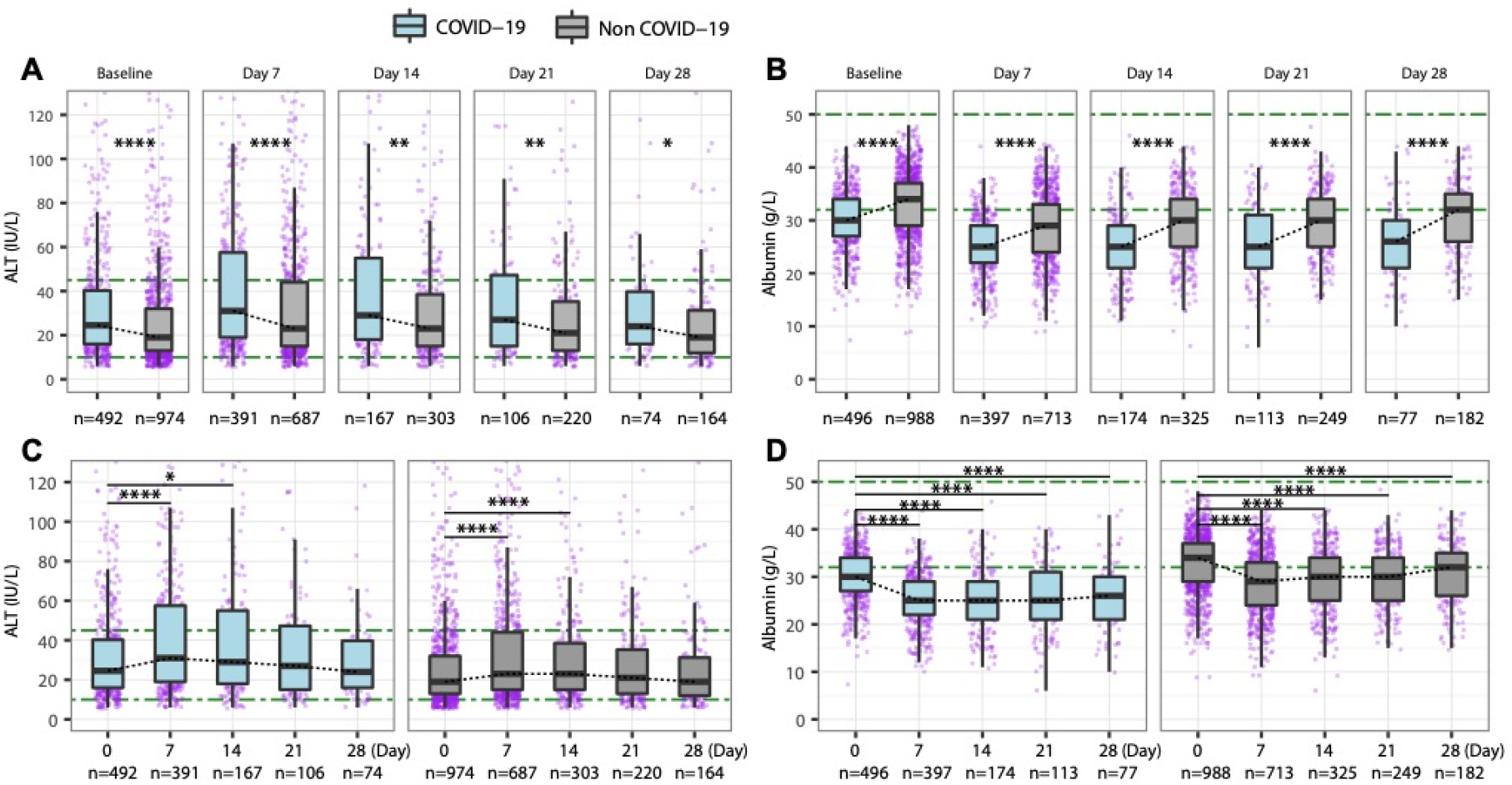
Comparison of ALT and albumin between hospitalised adults with COVID-19 and non-COVID-19 patients matched for age, gender, ethnicity and pre-existing comorbidities (**A**) ALT comparison at baseline, and 7, 14, 21, 28 days; (**B**) Albumin comparison at baseline, and 7, 14, 21, 28 days; (**C**) ALT changes over time in COVID-19 and non-COVID-19 groups; (**D**) Albumin changes over time in COVID-19 and non-COVID-19 groups. *ALT, Alanine transaminase. Green dash-dotted lines indicate the lower limits of normal and the upper limits of normal. * p-value <0.05, ** p-value <0.01, *** p-value <0.001, **** p-value <0.0001*.

### Demographics, comorbidities and liver biochemical characteristics associated with mortality in COVID-19 patients

In the COVID-19 group, patients who died were significantly older than those who survived (median age 82 years vs. 66 years respectively), significantly more likely to have pre-existing comorbidities including liver disease, DM, CHD and cancer (**Table 2)**, significantly more likely to have baseline hypoalbuminaemia (72.6% vs. 52.9%, p<0.001) and low nadir albumin during follow-up (96.2% vs. 72.7%, p<0.001, **Table 2**). In patients who died, baseline ALP was higher (p=0.007) and peak BR was more likely to be abnormal compared to those who survived (p=0.016, **Table S9**). Prescribed antiviral drug use was not different between groups (**Table S9**). Equivalent data for the non-COVID-19 group are provided in **Table S10**.

**Table 2.** Demographics, pre-existing comorbidities, baseline and peak/nadir liver biochemistry of COVID-19 patients who survived and died. For categorical variables, Fisher exact test was performed for comparison on cells with small counts (<5), otherwise Chi-square test was used. For continuous variables, Wilcoxon test was used for comparison due to non-normality. ^#^ BMI categories were reported based on WHO classification. ^†^ 82 vs. 11 in alive subgroup vs. died subgroup had not all liver biochemistry baseline data available. ^†^ 82 vs. 11 in alive subgroup vs. died subgroup missing baseline ALT. ^§^ 78 vs. 11 in alive subgroup vs. died subgroup missing baseline albumin. *ALT, Alanine transaminase; ALP, Alkaline phosphatase; CHD, Coronary heart disease; CKD, Chronic kidney disease; DM, Diabetes mellitus; HTN, Hypertension; IQR, Interquartile range; LLN, Lower limit of normal; ULN, Upper limit of normal*.

|  | **Survived (n=428)** | **Died (n=157)** | **p-value** |
| --- | --- | --- | --- |
| Gender = male, n(%) | 221 (51.6) | 91 (58.0) | 0.21 |
| Age at test (median [IQR]), years | 66 [54, 80] | 82 [75, 89] | **<0.001** |
| Age ≥ 75 years, n(%) | 158 (36.9) | 120 (76.4) | **<0.001** |
| **Ethnicity category, n(%)** |  |  |  |
| Asian | 29 (6.8) | 2 (1.3) | **0.015** |
| Black | 18 (4.2) | 2 (1.3) | 0.14 |
| Mixed and other | 17 (4.0) | 2 (1.3) | 0.17 |
| White | 287 (67.1) | 128 (81.5) | **0.001** |
| Not stated | 77 (18.0) | 23 (14.6) | 0.41 |
| BMI (median [IQR]), kg/m<sup>2</sup> | 27 [23.4, 31.3] | 25 [22, 30.2] | 0.076 |
| **BMI category<sup>#</sup>, n(%)** |  |  |  |
| <18.5 (underweight) | 10 (3.5) | 7 (6.7) | 0.28 |
| 18.5-24.9 (normal weight) | 98 (34.3) | 42 (40.0) | 0.35 |
| 25.0-29.9 (pre-obesity/overweight) | 87 (30.4) | 28 (26.7) | 0.55 |
| 30.0-34.9 (obesity class I) | 57 (19.9) | 20 (19.0) | 0.96 |
| 35.0-39.9 (obesity class II) | 21 (7.3) | 5 (4.8) | 0.49 |
| ≥40.0 (obesity class III) | 13 (4.5) | 3 (2.9) | 0.65 |
| Used invasive ventilation in ICU, n(%) | 41 (9.6) | 6 (3.8) | **0.036** |
| ICU admission, n(%) | 59 (13.8) | 12 (7.6) | 0.061 |
| **Pre-existing comorbidities, n(%)** |  |  |  |
| Liver disease | 6 (1.4) | 11 (7.0) | **0.001** |
| DM | 53 (12.4) | 35 (22.3) | **0.005** |
| HTN | 121 (28.3) | 58 (36.9) | 0.055 |
| CHD | 35 (8.2) | 24 (15.3) | **0.018** |
| CKD | 36 (8.4) | 20 (12.7) | 0.16 |
| Cancer | 24 (5.6) | 17 (10.8) | **0.045** |
| ≥1 liver biochemistry abnormal at baseline<sup>†</sup>, n(%) | 235 (67.9) | 122 (83.6) | **0.001** |
| ≥1 liver biochemistry abnormal at baseline (excl. albumin), n(%) | 123 (35.5) | 55 (37.7) | 0.73 |
| Baseline ALT (median [IQR]), IU/L | 25 [16, 43] | 23 [16, 36] | 0.11 |
| **Baseline ALT categories<sup>‡</sup>, n(%)** |  |  |  |
| normal | 267 (77.2) | 123 (84.2) | 0.099 |
| >1-2ULN | 49 (14.2) | 15 (10.3) | 0.31 |
| >2-3ULN | 16 (4.6) | 5 (3.4) | 0.72 |
| >3ULN | 14 (4.0) | 3 (2.1) | 0.40 |
| Baseline albumin (median [IQR]), g/L | 31 [28, 34] | 28 [25, 32] | **<0.001** |
| Baseline albumin (<LLN), n(%) | 185 (52.9) | 106 (72.6) | **<0.001** |
| ≥1 peak/nadir liver biochemistry abnormal, n(%) | 352 (82.2) | 153 (97.5) | **<0.001** |
| ≥1 peak liver biochemistry abnormal (excl. albumin), n(%) | 238 (55.6) | 91 (58.0) | 0.68 |
| Peak ALT (median [IQR]), IU/L | 37 [20, 71] | 32 [19, 52] | 0.07 |
| **Peak ALT categories, n(%)** |  |  |  |
| normal | 252 (58.9) | 111 (70.7) | **0.012** |
| >1-2ULN | 93 (21.7) | 22 (14.0) | **0.0496** |
| >2-3ULN | 28 (6.5) | 11 (7.0) | 0.99 |
| >3ULN | 55 (12.9) | 13 (8.3) | 0.17 |
| Nadir albumin (median [IQR]), g/L | 27 [22, 32] | 23 [19, 27] | **<0.001** |
| Nadir albumin (<LLN)<sup>§</sup>, n(%) | 311 (72.7) | 151 (96.2) | **<0.001** |

Survival curves and logrank tests (unadjusted for relevant confounders) within the COVID-19 group displayed that surviving probability over time was significantly lower in patients with hypoalbuminaemia at baseline than those with normal baseline albumin (**Figure 3A**), a higher surviving probability in patients with elevated baseline ALT as compared to normal baseline ALT (**Figure 3B**), while elevated ALP or bilirubin at baseline were not significantly associated with a lower survival probability over time (**Figure 3C-D**). In addition, for the subset of COVID-19 patients who had prothrombin time (PT) and international normalised ratio (INR), elevation of these parameters at baseline were significantly associated with a lower survival probability (**Figure S3**).

**Figure 3.**
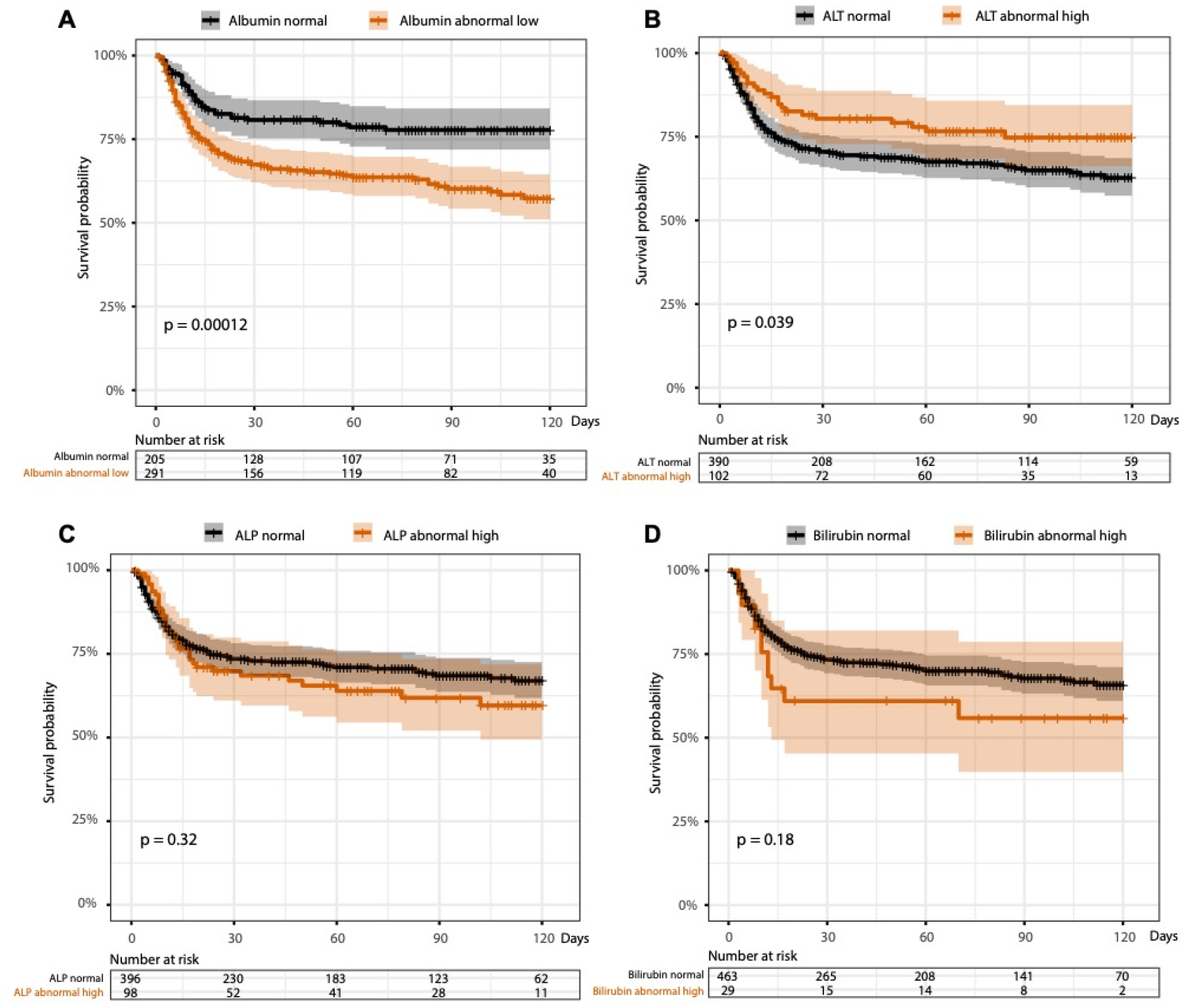
Survival K-M curves stratified by baseline liver biochemistry at the time of testing positive for SARS-CoV-2. (**A**) normal and low baseline albumin; (**B**) normal and elevated baseline ALT; (**C**) normal and elevated baseline ALP; (**D**) normal and elevated baseline bilirubin. *K-M, Kaplan-Meier. p-values were based on the logrank test. One patient has missing death date*.

In multivariate analysis (fully adjusted for demographics, comorbidities, and prescribed drug use before baseline) for the COVID-19 group, those with hypoalbuminaemia at baseline had a 1.83-fold-increased hazards of death compared to those with normal baseline albumin (adjusted HR 1.83, 95% CI 1.25-2.67), those aged ≥75 years had a ~4-fold-increased hazards of death compared to those aged <75 years (adjusted HR 3.96, 95% CI 2.59-6.04), and those with pre-existing liver disease had a ~3-fold-increased hazards of death than those without pre-existing liver disease (adjust HR 3.37, 95% CI 1.58-7.16) (**Table 3**). More specifically, a one unit (1 g/L) decrease in albumin at baseline was associated with a 5% increase in hazards of death (adjusted HR 1.05, 95%CI 1.02-1.09), while age increased by 10 years was associated with 82% increase in hazards of death (adjusted HR 1.82, 95% CI 1.56-2.13) (**Table S11**). Similarly, a one unit decrease in nadir albumin during follow-up was associated with a 7% increase in hazards of death (adjusted HR 1.07, 95% CI 1.04-1.10) (**Table S12**). In the COVID-19 group baseline albumin was significantly negatively correlated with age in those that survived (R=-0.264, p<0.0001) but not in those who died (R=-0.027, p=0.75) (**Figure S4**).

**Table 3.** Univariate and multivariate Cox proportional-hazards models investigating associations of baseline liver biochemistry derangement with death among adults with confirmed COVID-19. Analysis were performed on the 492 COVID-19 patients who had baseline liver biochemistry data (survived vs. died: 346 vs. 146); the cut off of age was based on the median age in the cohort; variables (except for demographics and comorbidities) with p<0.1 in univariate analysis were included for multivariate analysis; in multivariate analysis, HRs were fully adjusted for drugs use before baseline (including antiviral drugs, antibiotics, anticoagulants, acetaminophen, immunosuppressants, statins) to reduce confounding effects. *ALT, Alanine transaminase; ALP, Alkaline phosphatase; CHD, Coronary heart disease; CKD, Chronic kidney disease; DM, Diabetes mellitus; HTN, Hypertension; HR, Hazards ratio; LLN, lower limit of normal; ULN, upper limit of normal*.

| Variables | Univariate analysis |  | Multivariate analysis |  |
| --- | --- | --- | --- | --- |
|  | Crude HR (95% CIs) | p-value | Adjusted HR (95% CIs) | p-value |
| Age $\geq 75$ years | 4.39 (3.03-6.36) | **<0.001** | **3.96 (2.59-6.04)** | **<0.001** |
| Gender (male) | 1.24 (0.91-1.71) | 0.18 | 1.21 (0.86-1.71) | 0.28 |
| Ethnicity (white) | 1.87 (1.25-2.8) | **0.002** | 1.13 (0.71-1.8) | 0.61 |
| Baseline ALT ($>ULN$) | 0.63 (0.4-0.98) | **0.04** | 0.86 (0.53-1.38) | 0.53 |
| Baseline ALP ($>ULN$) | 1.21 (0.83-1.78) | 0.33 | | |
| Baseline bilirubin ($>ULN$) | 1.5 (0.83-2.7) | 0.18 | | |
| Baseline albumin ($<LLN$) | 2.01 (1.4-2.9) | **<0.001** | **1.83 (1.25-2.67)** | **0.002** |
| Pre-existing liver disease | 2.66 (1.44-4.9) | **0.002** | **3.37 (1.58-7.16)** | **0.002** |
| Pre-existing DM | 1.7 (1.16-2.48) | **0.006** | 1.22 (0.77-1.93) | 0.39 |
| Pre-existing HTN | 1.33 (0.96-1.84) | 0.085 | 0.7 (0.44-1.11) | 0.13 |
| Pre-existing CHD | 1.72 (1.11-2.65) | **0.015** | 1.16 (0.67-2.02) | 0.59 |
| Pre-existing CKD | 1.34 (0.83-2.17) | 0.23 | 0.84 (0.46-1.51) | 0.56 |
| Pre-existing Cancer | 1.71 (1.04-2.84) | **0.036** | 1.06 (0.58-1.93) | 0.85 |

For the subset of COVID-19 patients who had BMI data (survived vs. died, 246 vs. 96), HRs did not change materially after additional adjustment for BMI in the multivariate analysis (**Table S13**). Baseline albumin was weakly correlated with BMI both in those who died and survived (R=0.08, p=0.42 vs. R=0.099, p=0.12, respectively) (**Figure S5**).

### Longitudinal assessment of liver biochemistry patterns in patients who died with COVID-19, compared to those who survived

Within the COVID-19 group, patients who died during follow-up had significantly lower median values of albumin at baseline, 7 and 14 days after a positive SARS-CoV-2 RT-PCR, compared to the patients who survived (all p<0.001) (**Figure 4A**). There was no significant difference in ALT at any time point other than 7 days between those who died and survived (**Figure 4B**), while ALP was higher at baseline and 28 days (both p<0.05) and BR was higher at 7 days (p<0.05) (**Figure 4C-D**).

**Figure 4.**
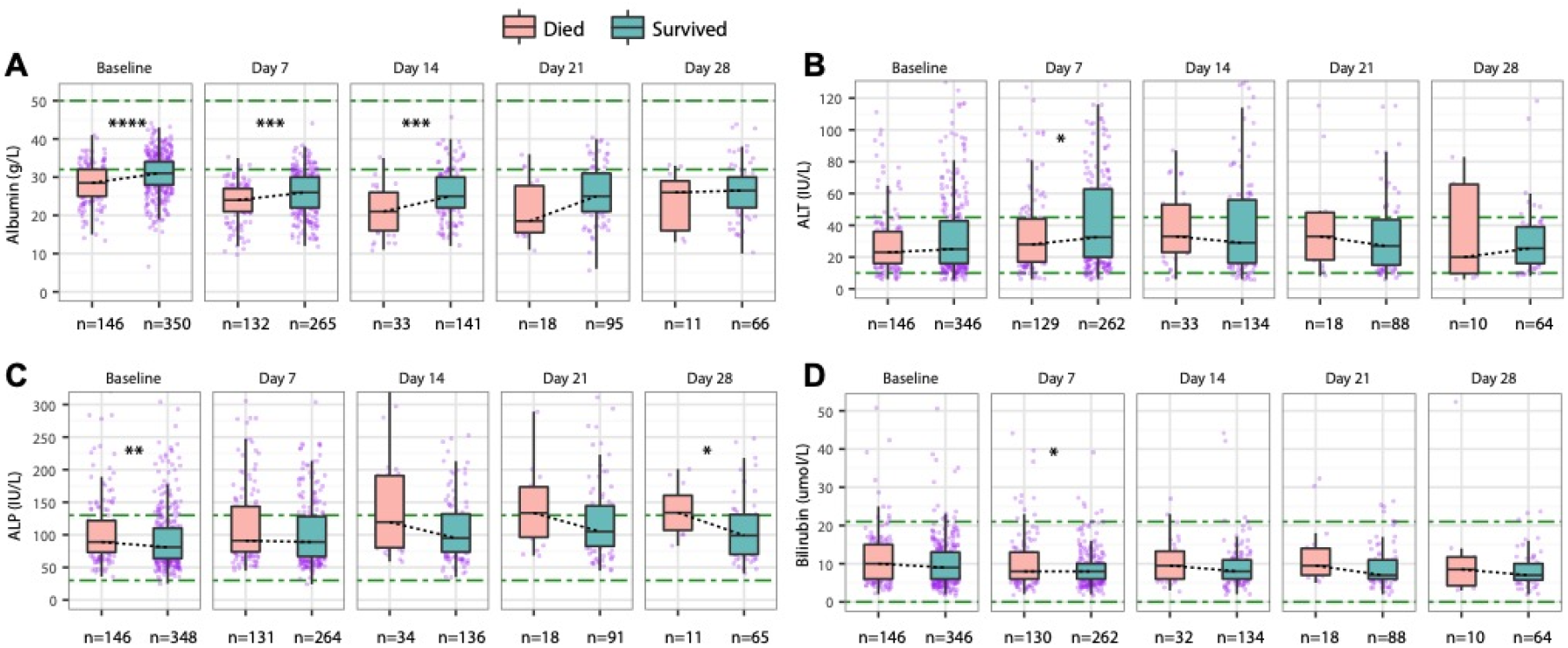
Comparison of liver biochemistry between patients with COVID-19 who died during follow-up and who survived to the end of follow-up. (**A**) Albumin comparison at baseline at baseline, 7, 14, 21, 28 days; (**B**) ALT comparison at baseline, 7, 14, 21, 28 days; (**C**) ALP comparison at baseline at baseline, 7, 14, 21, 28 days; (**D**) Bilirubin comparison at baseline at baseline, 7, 14, 21, 28 days. *ALT, Alanine transaminase; ALP, Alkaline phosphatase. Green dash-dotted lines indicate the lower limits of normal and the upper limits of normal. * p-value <0.05, ** p-value <0.01, *** p-value <0.001, ****p-value <0.0001*.

In the COVID-19 group, ALT increased during the first two weeks and remained elevated during follow-up in patients who died, whilst in patients who survived ALT decreased from 7 days onward with a trend to normalisation (**Figure 5A-B**). ALP increased over time in both subgroups, with a greater increase in those who died (**Figure 5C-D**). BR decreased throughout follow-up in both subgroups (**Figure 5E-F**). Albumin decreased significantly in both groups during the first 7 days and continued to decline in patients who died, but in patients who survived had an upward trend especially for those had been followed up longer than one month (**Figure 5G-H**).

**Figure 5.**
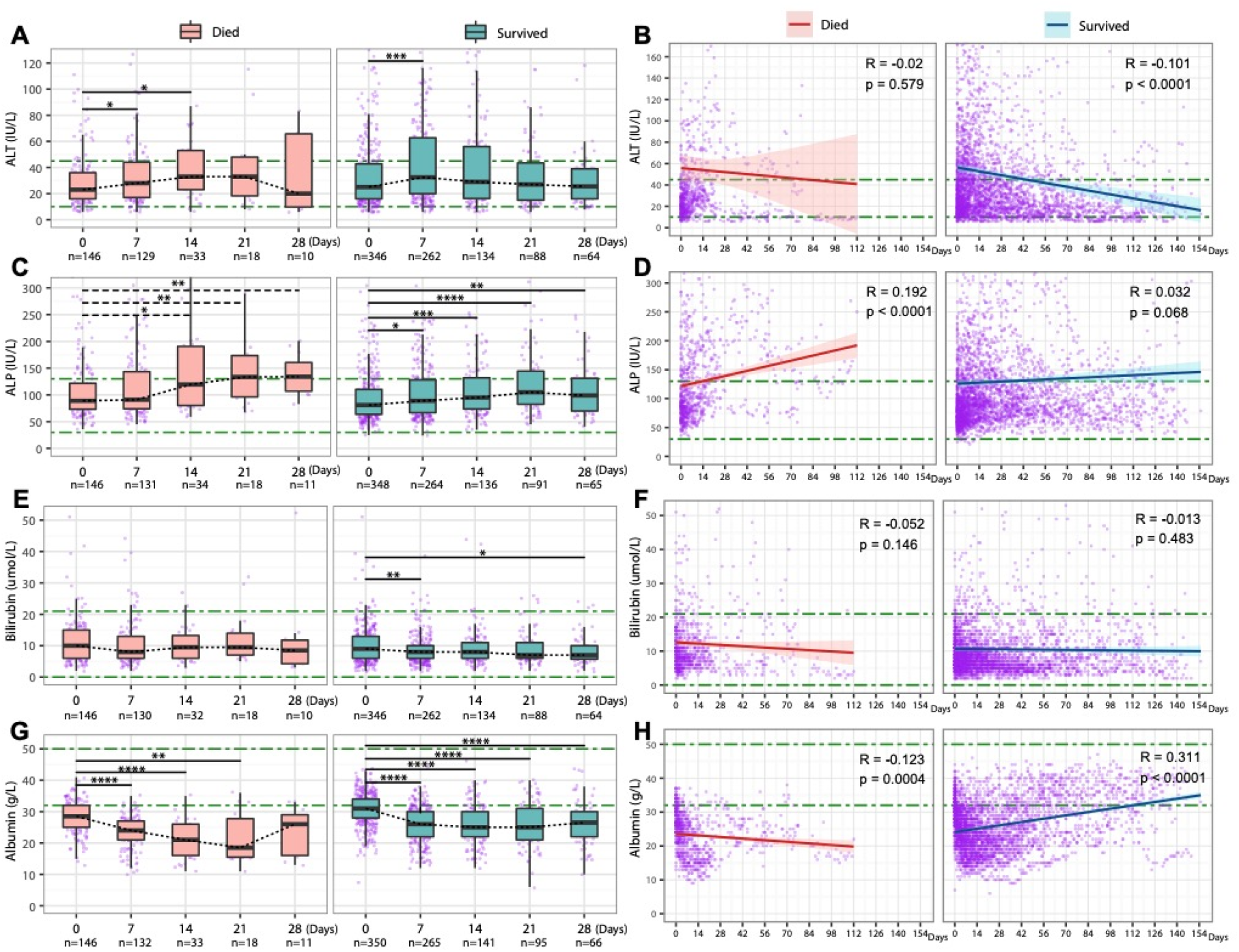
Longitudinal changes of liver biochemistry over time of COVID-19 patients stratified by death during follow-up. (**A**) ALT at baseline, 7, 14, 21, 28 days; (**B**) Changing trend over time (112 vs. 155 days) of ALT by linear regression line fitting with 95% CI; (**C**) ALP at baseline at baseline, 7, 14, 21, 28 days; (**D**) Changing trend over time (112 vs. 155 days) of ALP by linear regression line fitting with 95% CI; (**E**) Bilirubin at baseline at baseline, 7, 14, 21, 28 days; (**F**) Changing trend over time (112 vs. 155 days) of bilirubin by linear regression line fitting with 95% CI; (**G**) Albumin at baseline at baseline, 7, 14, 21, 28 days; (**H**) Changing trend over time (112 vs. 155 days) of albumin by linear regression line fitting with 95% CI. *ALT, Alanine transaminase; ALP, Alkaline phosphatase; CI, Confidence interval. Green dash-dotted lines indicate the lower limits of normal and the upper limits of normal. * p-value <0.05, ** p-value <0.01, *** p-value <0.001, **** p-value <0.0001. R represents Pearson’s correlation coefficient and p indicates linear regression significance*.

Among COVID-19 patients who had ≥2 longitudinal data points for ALT, ALP, BR and albumin, 468 had at least one liver biochemistry derangement during follow-up. Among them 26.9% (126/468) had normalised by the end of follow-up, while the remaining 73.1% still had at ≥1 abnormal liver biochemistry at the end of follow-up. The information of each liver biochemistry recovery were provided in **Table S14**.

## Discussion

### Novelty and key findings

To our knowledge this is the first study to: i) comprehensively conduct longitudinal analyses of liver biochemistry patterns over time in COVID-19 patients compared to a matched general patient cohort, and ii) analyse longitudinal liver biochemistry patterns in COVID-19 patients who have died and survived. We have gathered a large cohort through an automatic approach, and account for missing data in the analysis.

The COVID-19 group exhibited ~2-fold higher death rate, with significantly lower survival probability compared to the non-COVID-19 group. Among the COVID-19 group, a higher proportion of patients had at least one abnormal liver biochemistry (ALT, ALP, BR or albumin) at baseline and during follow-up, compared to the matched non-COVID-19 group. Patients with COVID-19 who died showed a decline in albumin and a greater increase in ALP over time compared to those who survived, and baseline hypoalbuminaemia was a significant predictor of death in COVID-19 patients on multivariate analysis with adjusting for relevant confounders.

### Comparison to previous studies

In our study, rates of baseline and peak ALT derangement between COVID-19 and non-COVID-19 groups were significantly different, consistent with findings from a large USA cohort (8). The increase in ALT between baseline and at 14 days follow-up in the COVID-19 group is also consistent with previous research (6).

Patients with pre-existing liver disease were found to have an increased risk of mortality in COVID-19 which is consistent with the findings of previous studies (19, 20) however there is only a small number of patients with the pre-existing liver disease in our study. Because some COVID-19 patients in our cohort were admitted to hospital for non-COVID-19 we analysed baseline liver biochemistry at the time of the SARS-CoV-2 RT-PCR rather than date of admission. This may partially explain differences between our study and previous studies which analysed liver biochemistry measured on hospital admission (3-6). Variable patterns of drug use between cohorts may also account for differences.

Although baseline hypoalbuminaemia was significantly associated with hazards of death in COVID-19, ALT, ALP, and BR were not. Recent findings from two multi-centre studies support the prognostic association of albumin with death in COVID-19 (16, 21). However, other studies aiming to explore whether liver biochemistry derangement is associated with death in COVID-19 did not investigate albumin (6-8). Interestingly, a previous study found that hypoalbuminemia is a strong predictor of 30-day all-cause mortality in acutely admitted medical patients (22). As albumin is a cheap and widely available test, it can be usefully employed as a prognostic biomarker.

Due to variable population settings in previous studies (e.g., demographics, comorbidities), it is important to understand the populations in which prognostic models are developed (12) to ensure such models are externally valid. For example, a study from the US (8) reported a positive association of ALT >5 times ULN with death, however we were unable to replicate this investigation as only a small number of participants had ALT elevated to this level; furthermore, the prevalence of comorbid disease and BMI >35kg/m^2^ in the US study population was much higher compared to our cohort. Similarly, a large primary care cohort study (23) reported that BMI >40kg/m^2^ was associated with an increased risk of COVID-19-related death, but we were underpowered to replicate this analysis.

### COVID-19 and liver biochemistry derangement

Although derangements in liver biochemistry are common in COVID-19 patients, the reasons for the liver injury remain unclear, but may include direct viral damage, drug-induced liver injury, hypoxia, immune-mediated injury, sepsis, or cytokine release (24, 25). Angiotensin-converting enzyme 2 (ACE2), a functional receptor for SARS-CoV-2 (26) is found abundantly in the gastrointestinal tract and liver, in addition to presenting in alveolar type 2 cells (the major SARS-CoV-2 targeting cell type in lung). A recent study observed a higher expression of ACE2 in cholangiocytes (59.7% of cells) compared to hepatocytes (2.6%) (27). Given the hepatic distribution of the ACE2 receptor, SARS-CoV-2 may well cause damage of both bile ducts and liver (28, 29). Alternatively, the liver may be a bystander, with deranged liver biochemistry reflecting systemic disease (30).

### Caveats and limitations

Routinely collected liver biochemistry is not consistent between settings, and therefore the definitions of liver biochemistry derangement may vary across studies. Although AST and GGT have been investigated in previous studies, these parameters are not available for our population. We recognise that analysis can be influenced by missing data, but we report missing values and investigating peak/nadir values as well as baseline. We also undertook sensitivity analysis in a subset of patients with complete BMI measurements to investigate its association with death. Our cohort in the South East of the UK may not be representative of populations elsewhere, especially in terms of ethnic diversity, so caution should be applied in extrapolation of results.

### Future studies

Further longitudinal studies of COVID-19 outcomes in diverse patient groups, including those with pre-existing liver disease are needed. The NIHR HIC program will continue to benefit the field of COVID-19 research, as data accumulated for further large teaching hospitals can be used to expand this analysis, resulting in a more generalizable study population and increased statistical power.

### Conclusion

Liver biochemistry derangement is common in COVID-19 patients at the time of a SARS-CoV-2 RT-PCR test and during the clinical course of disease. Baseline and nadir albumin are valuable prognostic factors that are associated with death in COVID-19 patients.

## Data Availability

N/A

## Author Contributions

T.W, D.A.S, C.C., P.C.M and E.B contributed equally to this work. T.W, D.A.S and C.C performed the data analysis and wrote the manuscript and were supervised by E.B, P.C.M and J.D. Infrastructure development and data collection was performed by S.H, H.S., K.A.V, K.W, J.D. All authors reviewed the manuscript and provided significant edits to the manuscript.

## Supplementary Tables

**Table S1.** Normal reference and units of blood tests used in this study, based on reference ranges set by the hospital biochemistry lab.

| Categories | Test name | Normal reference | Units |
| --- | --- | --- | --- |
| Liver function | Alanine transaminase (ALT) | 10-45 | IU/L |
|  | Alkaline phosphatase (ALP) | 30-130 | IU/L |
|  | Bilirubin | 0-21 | umol/L |
|  | Albumin | 32-50 | g/L |
| Blood clotting tests | Prothrombin time (PT) | 9.0-12.0 | seconds |
|  | Activated partial thromboplastin time (APTT) | 20.0-30.0 | seconds |
|  | International normalised ratio (INR) | 0.8-1.2 | ratio |
| Renal function tests | Creatinine | 64-104 | umol/L |
|  | Urea | 3.0-9.2 | mmol/L |
|  | Estimated Glomerular Filtration Rate (eGFR) | ≥90 | ml/min/1.73m<sup>2</sup> |
| Other tests | C-reactive protein (CRP) | 0-5 | mg/L |
|  | Platelets | 150-400 | x10<sup>9</sup>/L |
|  | Lymphocytes | 1.0-4.0 | x10<sup>9</sup>/L |

**Table S2.** Historical diagnostic ICD codes used to identify pre-existing comorbidities of interest in this study.

| Pre-existing comorbidities | ICD code categories (retrieved) | notes |
| --- | --- | --- |
| Liver disease | B18 category | Chronic viral hepatitis |
|  | K70-K77 categories | Diseases of liver<br>• K70 category: Alcoholic liver disease<br>• K71 category: Toxic liver disease<br>• K72 category: Hepatic failure, not elsewhere classified<br>• K73 category: Chronic hepatitis, not elsewhere classified<br>• K74 category: Fibrosis and cirrhosis of liver<br>• K75 category: Other inflammatory liver diseases<br>• K76 category: Other diseases |
|  |  | of liver<br>K77 category: Liver disorders in diseases classified elsewhere |
| DM | E10-E14 categories | Diabetes mellitus |
| HTN | I10 category | Essential (primary) hypertension |
| CHD | I20-I25 categories | Coronary heart disease, *also called* ischaemic heart disease |
| CKD | N18 category | Chronic kidney disease |
| Cancer | C00-C97 categories | Malignant neoplasms |

**Table S3.**
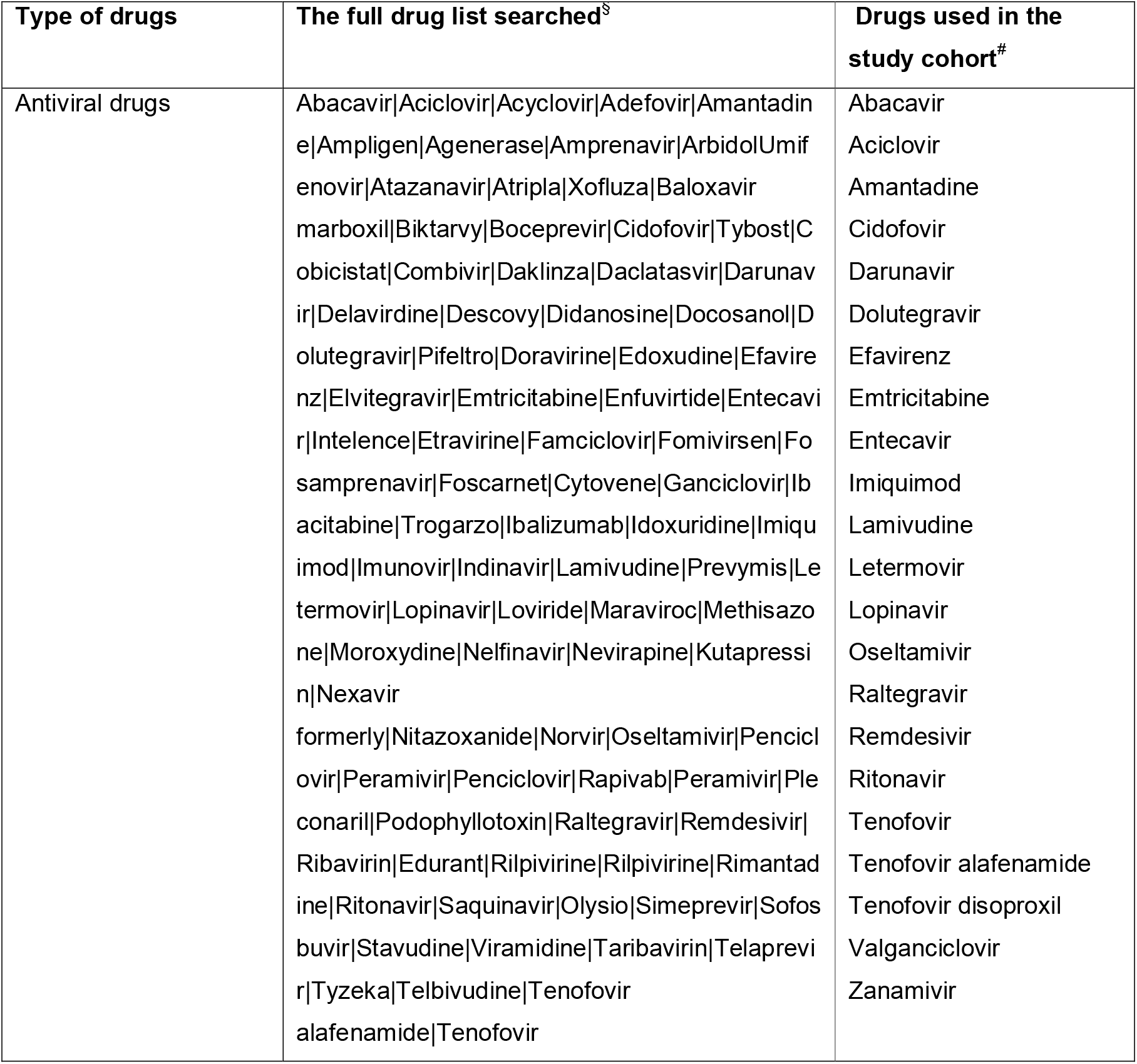
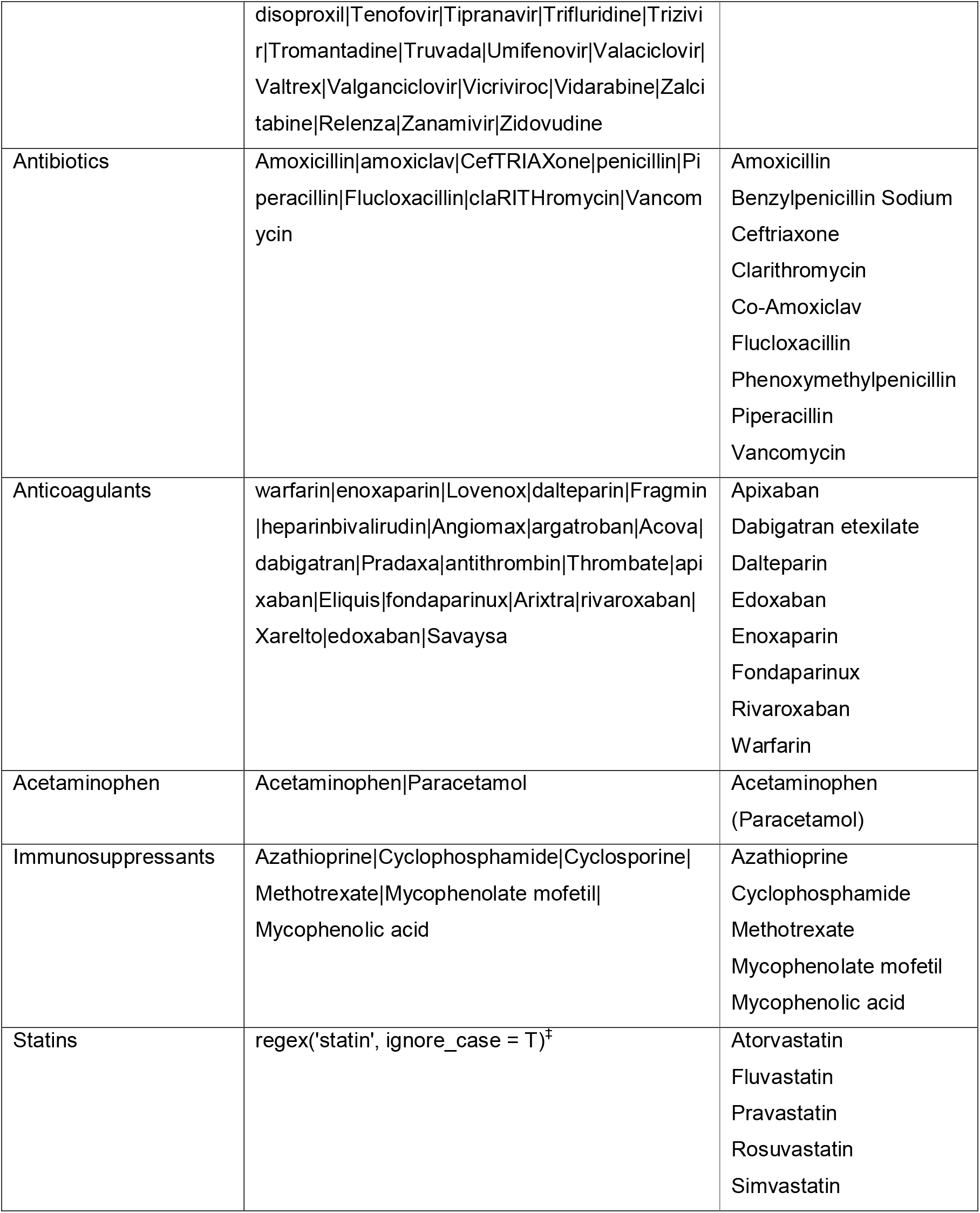
Detailed information for the full list of drugs searched for in this cohort, and drugs used in the study cohort. § Regular expressions were used to retrieve the drug information. *Statins were searched using a regular expression including ‘statin’. ^#^The drugs used as part of clinical trials were not recorded in the dataset.

**Table S4.** Demographics and comorbidity characteristics of patients with and without COVID-19 before and after propensity score matching.

|  | Before matching |  |  | After matching |  |  |
| --- | --- | --- | --- | --- | --- | --- |
| Variables used for matching | COVID-19 Positive (n=585) | COVID-19 Negative (n=5726) | p-value | COVID-19 Positive (n=585) | COVID-19 Negative (n=1165) | p-value |
| Gender = male (%) | 312 (53.3) | 2901 (50.7) | 0.235 | 312 (53.3) | 629 (54.0) | 0.834 |
| Age at test (median [IQR]) | 73 [57, 84] | 68 [53, 81] | **<0.001** | 73 [57, 84] | 73 [58, 83] | 0.755 |
| Ethnicity category (%) |  |  | **<0.001** |  |  | 0.911 |
| Asian | 31 (5.3) | 166 (2.9) |  | 31 (5.3) | 62 (5.3) |  |
| Black | 20 (3.4) | 65 (1.1) |  | 20 (3.4) | 35 (3.0) |  |
| Mixed | 10 (1.7) | 41 (0.7) |  | 10 (1.7) | 22 (1.9) |  |
| Not stated | 100 (17.1) | 1062 (18.5) |  | 100 (17.1) | 219 (18.8) |  |
| Other | 9 (1.5) | 43 (0.8) |  | 9 (1.5) | 23 (2.0) |  |
| White | 415 (70.9) | 4349 (76.0) |  | 415 (70.9) | 804 (69.0) |  |
| **Pre-existing comorbidities, n (%)** |  |  |  |  |  |  |
| Liver disease | 17 (2.9) | 210 (3.7) | 0.409 | 17 (2.9) | 33 (2.8) | 1 |
| DM | 88 (15.0) | 572 (10.0) | **<0.001** | 88 (15.0) | 175 (15.0) | 1 |
| HTN | 179 (30.6) | 1100 (19.2) | **<0.001** | 179 (30.6) | 349 (30.0) | 0.825 |
| CHD | 59 (10.1) | 441 (7.7) | 0.051 | 59 (10.1) | 113 (9.7) | 0.864 |
| CKD | 56 (9.6) | 400 (7.0) | **0.027** | 56 (9.6) | 107 (9.2) | 0.86 |
| Cancer | 41 (7.0) | 628 (11.0) | **0.004** | 41 (7.0) | 88 (7.6) | 0.753 |

**Table S5.** Distribution of specialities under which patients in COVID-19 and non-COVID-19 groups were admitted to hospital.

|  | COVID-19 group (n=585) |  | non-COVID-19 group (n=1165) |  |
| --- | --- | --- | --- | --- |
| Specialty | count | percentage | count | percentage |
| General Internal Medicine | 213 | 36.40% | 267 | 22.90% |
| Geriatric Medicine | 115 | 19.70% | 87 | 7.50% |
| Infectious Diseases | 25 | 4.30% | 48 | 4.10% |
| Respiratory Medicine | 23 | 3.90% | 25 | 2.10% |
| Emergency Medicine | 22 | 3.80% | 44 | 3.80% |
| Gastroenterology | 21 | 3.60% | 42 | 3.60% |
| General Surgery | 19 | 3.20% | 213 | 18.30% |
| Trauma and Orthopaedics | 15 | 2.60% | 77 | 6.60% |
| Renal Medicine | 13 | 2.20% | 28 | 2.40% |
| Endocrinology and Diabetes | 12 | 2.10% | 3 | 0.30% |
| Clinical Haematology | 10 | 1.70% | 21 | 1.80% |
| Cardiology | 8 | 1.40% | 43 | 3.70% |
| Clinical Pharmacology | 4 | 0.70% | 2 | 0.20% |
| Medical Oncology | 4 | 0.70% | 10 | 0.90% |
| Urology | 3 | 0.50% | 32 | 2.70% |
| Neurosurgery | 3 | 0.50% | 26 | 2.20% |
| Cardiothoracic Surgery | 3 | 0.50% | 12 | 1.00% |
| Plastic Surgery | 2 | 0.30% | 4 | 0.30% |
| Neurology | 2 | 0.30% | 9 | 0.80% |
| Gynaecology | 2 | 0.30% | 13 | 1.10% |
| Ophthalmology | 1 | 0.20% | 4 | 0.30% |
| Oral and Maxillofacial Surgery | 1 | 0.20% | 4 | 0.30% |
| Intensive Care Medicine | 1 | 0.20% | 7 | 0.60% |
| Palliative Medicine | 1 | 0.20% | 7 | 0.60% |
| Rheumatology | 1 | 0.20% | 2 | 0.20% |
| Clinical Oncology | 1 | 0.20% | 7 | 0.60% |
| Ear Nose and Throat |  |  | 5 | 0.40% |
| Radiology |  |  | 5 | 0.40% |
| Rehabilitation Medicine |  |  | 3 | 0.30% |
| Anaesthetics |  |  | 1 | 0.10% |
| Dermatology |  |  | 1 | 0.10% |
| Medical Microbiology |  |  | 1 | 0.10% |
| Midwifery |  |  | 1 | 0.10% |
| Obstetrics |  |  | 1 | 0.10% |
| Unknown | 60 | 10.30% | 110 | 9.40% |

**Table S6.** Drug use before and after baseline in COVID-19 and non-COVID-19 groups.

| Type of drugs | Drug use before baseline |  |  | Drug use after baseline |  |  |
| --- | --- | --- | --- | --- | --- | --- |
|  | COVID-19 group (n=585) | non COVID-19 group (n=1165) | p-value | COVID-19 group (n=585) | non COVID-19 group (n=1165) | p-value |
| Antiviral drugs use, n (%) | 20 (3.4) | 31 (2.7) | 0.460 | 44 (7.5) | 67 (5.8) | 0.184 |
| Antibiotics use, n (%) | 215 (36.8) | 275 (23.6) | **<0.001** | 424 (72.5) | 718 (61.6) | **<0.001** |
| Anticoagulants use, n (%) | 233 (39.8) | 344 (29.5) | **<0.001** | 462 (79.0) | 873 (74.9) | 0.070 |
| Acetaminophen use, n (%) | 258 (44.1) | 370 (31.8) | **<0.001** | 448 (76.6) | 896 (76.9) | 0.925 |
| Immunosuppressants use, n (%) | 13 (2.2) | 21 (1.8) | 0.677 | 17 (2.9) | 46 (3.9) | 0.333 |
| Statins use, n (%) | 128 (21.9) | 193 (16.6) | **0.008** | 201 (34.4) | 445 (38.2) | 0.129 |

**Table S7.** Patterns of missing data of liver biochemistry at baseline in COVID-19 and non-COVID-19 groups.

|  |  | COVID-19 group |  | non-COVID-19 group |  |
| --- | --- | --- | --- | --- | --- |
|  |  | No missing baseline data on all the four liver biochemistry (n=492) | Has missing baseline data on liver biochemistry (n=93) | No missing baseline data on all the four liver biochemistry (n=974) | Has missing baseline data on liver biochemistry (n=191) |
| Stratified by peak liver biochemistry | Deranged | 454 (92.3) | 51 (54.8) | 762 (78.2) | 80 (41.9) |
|  | Normal | 38 (7.7) | 42 (45.2) | 212 (21.8) | 111 (58.1) |
| Stratified by death status | Died | 146 (29.7) | 11 (11.8) | 130 (13.3) | 9 (4.7) |
|  | Survived | 346 (70.3) | 82 (88.2) | 844 (86.7) | 182 (95.3) |

**Table S8.** Comparison of baseline and peak/nadir liver biochemistry in patients with and without COVID-19, stratified by various degrees of derangement. ^§^ Only 492 vs. 974 patients in COVID-19 group vs. non-COVID-19 group had data of all the liver biochemistry available at baseline. ^f^ For ALT, ALP, bilirubin, we investigated the peak values while nadir values for albumin. For categorical variables, Fisher exact test was performed for comparison on cells with small counts (<5), otherwise Chi-square test was used. For continuous variables, Wilcoxon test was used for comparison due to non-normality. *ALT, Alanine transaminase; ALP, Alkaline phosphatase;LLN, Lower limit of normal; ULN, Upper limit of normal*.

|  | Baseline liver biochemistry<sup>s</sup> |  |  | Peak/nadir liver biochemistry<sup>†</sup> |  |  |
| --- | --- | --- | --- | --- | --- | --- |
|  | COVID-19 group<br>n=492 | Non-COVID-19 group<br>n=974 | p-value | COVID-19 group<br>n=585 | non-COVID-19 group<br>n=1165 | p-value |
| **Number of deranged liver biochemistry, (%)** |  |  |  |  |  |  |
| 0 | 135 (27.4) | 433 (44.5) | **<0.001** | 80 (13.7) | 323 (27.7) | **<0.001** |
| 1 | 234 (47.6) | 337 (34.6) | **<0.001** | 212 (36.2) | 350 (30.0) | **0.01** |
| 2 | 92 (18.7) | 146 (15.0) | 0.081 | 169 (28.9) | 298 (25.6) | 0.16 |
| 3 | 25 (5.1) | 42 (4.3) | 0.59 | 92 (15.7) | 123 (10.6) | **0.002** |
| 4 | 6 (1.2) | 16 (1.6) | 0.69 | 32 (5.5) | 71 (6.1) | 0.68 |
| ALT (median [IQR]), IU/L | 25 [16, 40] | 19 [13, 32] | **<0.001** | 34 [20, 66] | 26 [17, 51] | **<0.001** |
| ALT, >ULN, n(%) | 102 (20.7) | 142 (14.6) | **0.004** | 222 (37.9) | 323 (27.7) | **<0.001** |
| **ALT (categories), n(%)** |  |  |  |  |  |  |
| normal | 390 (79.3) | 832 (85.4) | **0.004** | 363 (62.1) | 842 (72.3) | **<0.001** |
| >1-2ULN | 64 (13.0) | 88 (9.0) | **0.023** | 115 (19.7) | 184 (15.8) | 0.050 |
| >2-3ULN | 21 (4.3) | 20 (2.1) | **0.024** | 39 (6.7) | 49 (4.2) | **0.035** |
| >3ULN | 17 (3.5) | 34 (3.5) | 1 | 68 (11.6) | 90 (7.7) | **0.009** |
| ALP (median [IQR]), IU/L | 85 [66, 114] | 90 [71, 124] | **0.005** | 106 [79, 155] | 104 [78, 149] | 0.69 |
| ALP, >ULN, n(%) | 98 (19.8) | 213 (21.6) | 0.47 | 199 (34.0) | 382 (32.8) | 0.65 |
| **ALP (abnormal categories), n(%)** |  |  |  |  |  |  |
| normal | 396 (80.2) | 772 (78.4) | 0.47 | 386 (66.0) | 783 (67.2) | 0.65 |
| >1-2ULN | 82 (16.6) | 163 (16.5) | 1 | 146 (25.0) | 278 (23.9) | 0.66 |
| >2-3ULN | 9 (1.8) | 28 (2.8) | 0.31 | 32 (5.5) | 55 (4.7) | 0.57 |
| >3ULN | 7 (1.4) | 22 (2.2) | 0.39 | 21 (3.6) | 49 (4.2) | 0.62 |
| Bilirubin (median [IQR]), umol/L | 9 [6, 13] | 10 [7, 16] | **<0.001** | 11 [8, 16] | 12 [8, 17] | 0.20 |
| Bilirubin, >ULN, n(%) | 29 (5.9) | 127 (13.0) | **<0.001** | 71 (12.1) | 201 (17.3) | **0.007** |
| **Bilirubin (categories), n(%)** |  |  |  |  |  |  |
| normal | 463 (94.1) | 847 (87.0) | **<0.001** | 514 (87.9) | 964 (82.7) | **0.007** |
| >1-2ULN | 22 (4.5) | 89 (9.1) | **0.002** | 52 (8.9) | 135 (11.6) | 0.10 |
| >2-3ULN | 3 (0.6) | 17 (1.7) | 0.13 | 9 (1.5) | 29 (2.5) | 0.27 |
| >3ULN | 4 (0.8) | 21 (2.2) | 0.097 | 10 (1.7) | 37 (3.2) | 0.10 |
| Albumin (median [IQR]), g/L | 30 [27, 34] | 34 [29, 37] | **<0.001** | 26 [21, 31] | 29 [24, 35] | **<0.001** |
| Albumin, <LLN, n(%) | 291 (58.7) | 346 (35.0) | **<0.001** | 462 (79.0) | 693 (59.5) | **<0.001** |

**Table S9.** Characteristics of baseline and peak of ALP and bilirubin, and drug use of patients, stratified by death status in COVID-19 group. ^†^ 80 vs. 11 patients in alive subgroup vs. died subgroup had no data of ALP available at baseline. * 82 vs. 11 patients in alive subgroup vs. died subgroup had no data of bilirubin available at baseline. For categorical variables, Fisher exact test was performed for comparison on cells with small counts (<5), otherwise Chi-square test was used. For continuous variables, Wilcoxon test was used for comparison due to non-normality. *ALP, Alkaline phosphatase; IQR, Interquartile range; ULN, Upper limit of normal*.

|  | Survived (n=428) | Died (n=157) | p-value |
| --- | --- | --- | --- |
| Baseline ALP (median [IQR]), IU/L | 81 [64, 110] | 89 [73, 122] | **0.007** |
| **Baseline ALP categories<sup>†</sup>, n(%)** |  |  |  |
| Normal | 284 (81.6) | 112 (76.7) | 0.26 |
| >1-2ULN | 55 (15.8) | 27 (18.5) | 0.55 |
| >2-3ULN | 4 (1.1) | 5 (3.4) | 0.18 |
| >3ULN | 5 (1.4) | 2 (1.4) | 1 |
| Baseline bilirubin (median [IQR]), umol/L | 9 [6, 13] | 10 [6, 15] | 0.056 |
| **Baseline bilirubin categories<sup>‡</sup>, n(%)** |  |  |  |
| Normal | 329 (95.1) | 134 (91.8) | 0.23 |
| >1-2ULN | 13 (3.8) | 9 (6.2) | 0.35 |
| >2-3ULN | 2 (0.6) | 1 (0.7) | 1 |
| >3ULN | 2 (0.6) | 2 (1.4) | 0.73 |
| Peak ALP (median [IQR]), IU/L | 105 [77, 151] | 110 [84, 179] | 0.076 |
| **Peak ALP categories, n(%)** |  |  |  |
| Normal | 290 (67.8) | 96 (61.1) | 0.16 |
| >1-2ULN | 104 (24.3) | 42 (26.8) | **0.001** |
| >2-3ULN | 15 (3.5) | 17 (10.8) | 0.91 |
| >3ULN | 19 (4.4) | 2 (1.3) | **<0.001** |
| Peak bilirubin (median [IQR]), umol/L | 11 [8, 15] | 12 [8, 18] | 0.096 |
| **Peak bilirubin categories, n(%)** |  |  |  |
| Normal | 385 (90.0) | 129 (82.2) | **0.016** |
| >1-2ULN | 32 (7.5) | 20 (12.7) | 0.069 |
| >2-3ULN | 5 (1.2) | 4 (2.5) | 0.41 |
| >3ULN | 6 (1.4) | 4 (2.5) | 0.56 |
| **Drug use, n(%)** |  |  |  |
| Antiviral drugs use (before baseline) | 13 (3.0) | 7 (4.5) | 0.442 |
| Antiviral drugs use (during follow-up) | 35 (8.2) | 9 (5.7) | 0.379 |
| Antibiotics use (before baseline) | 145 (33.9) | 70 (44.6) | **0.022** |
| Antibiotics use (during follow-up) | 287 (67.1) | 137 (87.3) | **<0.001** |
| Anticoagulants use (before baseline) | 154 (36.0) | 79 (50.3) | **0.002** |
| Anticoagulants use (during follow-up) | 335 (78.3) | 127 (80.9) | 0.565 |
| Acetaminophen use (before baseline) | 178 (41.6) | 80 (51.0) | 0.054 |
| Acetaminophen use (during follow-up) | 317 (74.1) | 131 (83.4) | **0.024** |
| Immunosuppressants use (before baseline) | 12 (2.8) | 1 (0.6) | 0.202 |
| Immunosuppressants use (during follow-up) | 14 (3.3) | 3 (1.9) | 0.579 |
| Statins use (before baseline) | 85 (19.9) | 43 (27.4) | 0.066 |
| Statins use (during follow-up) | 145 (33.9) | 56 (35.7) | 0.76 |

**Table S10.**
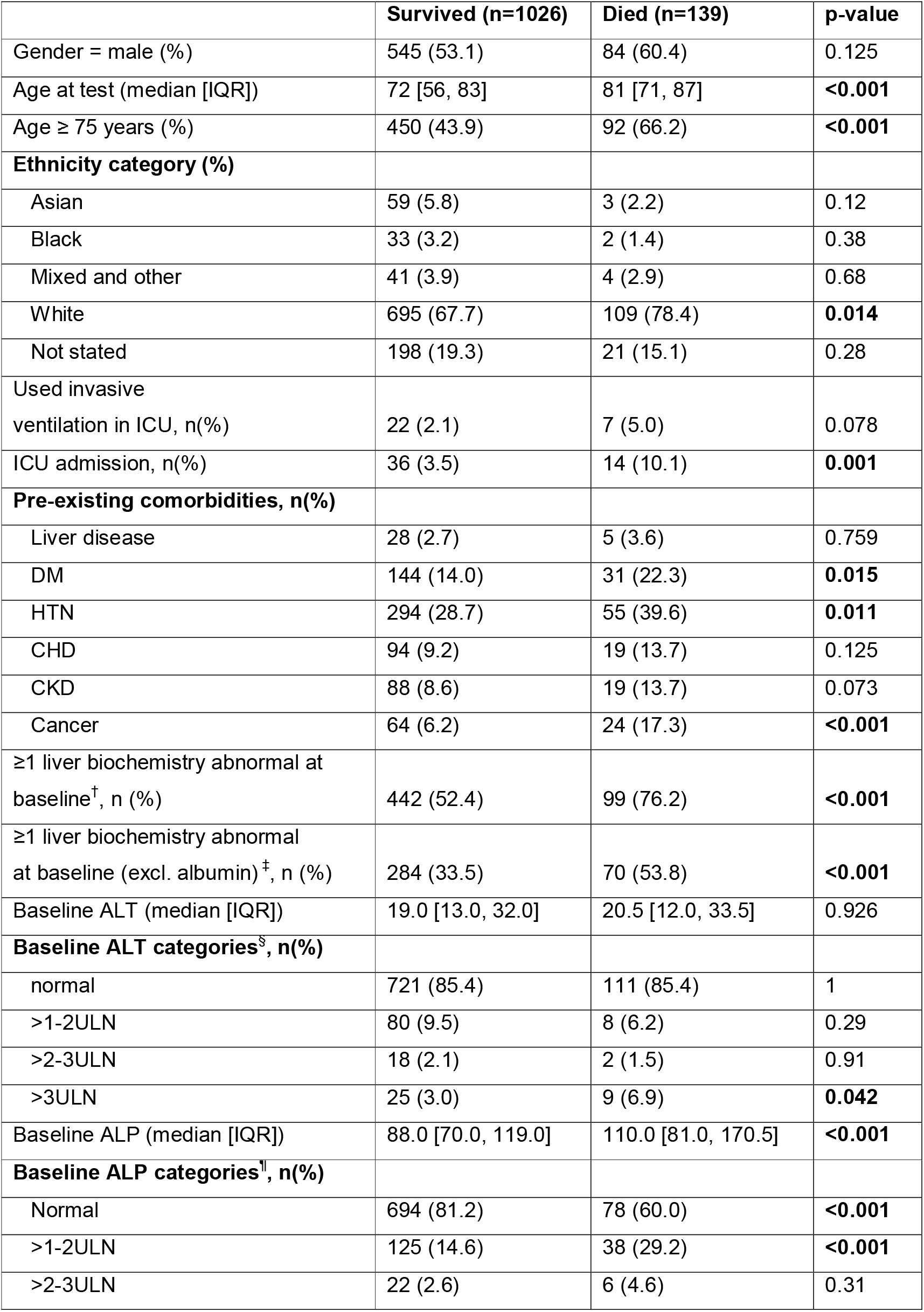
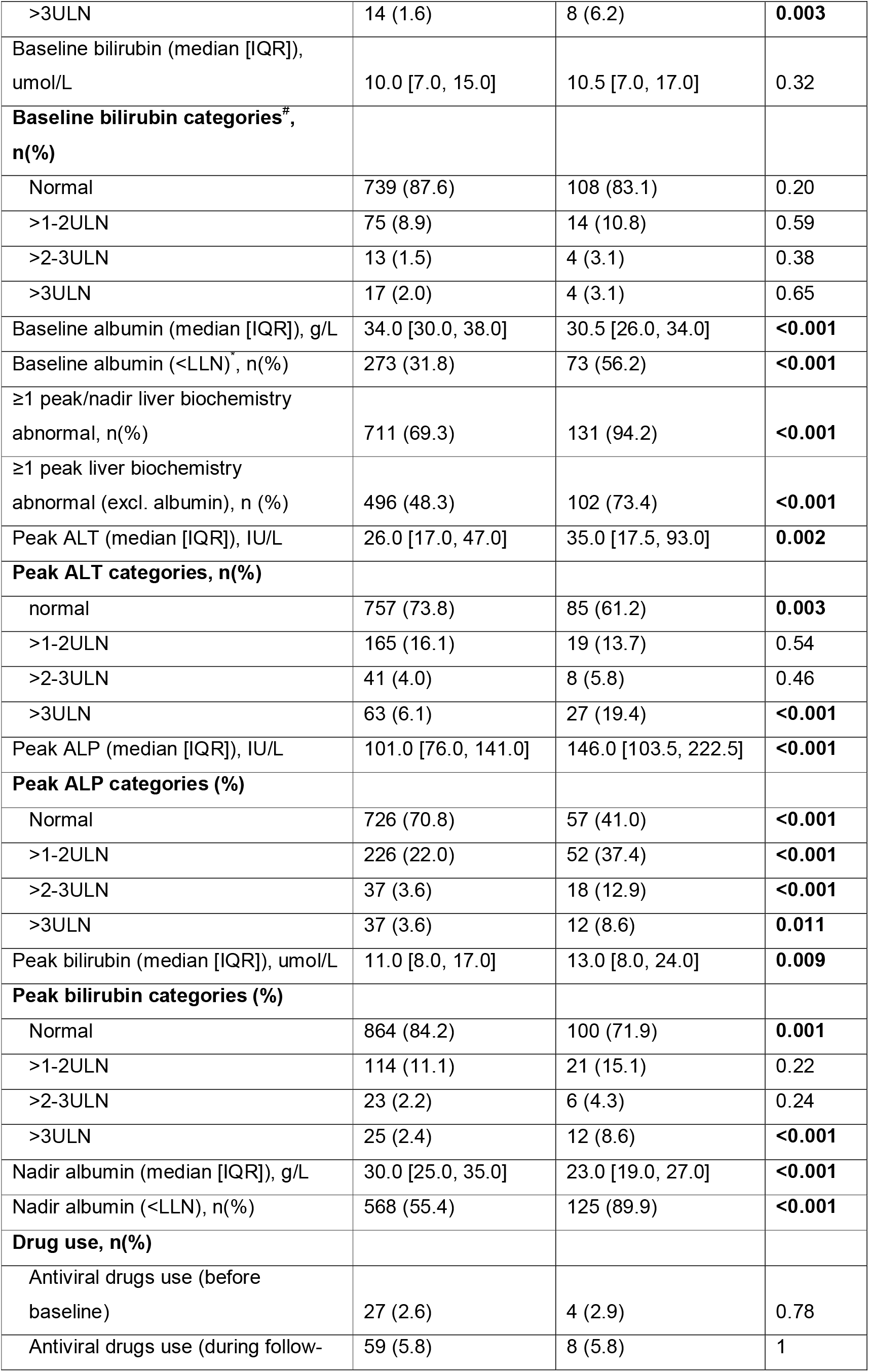
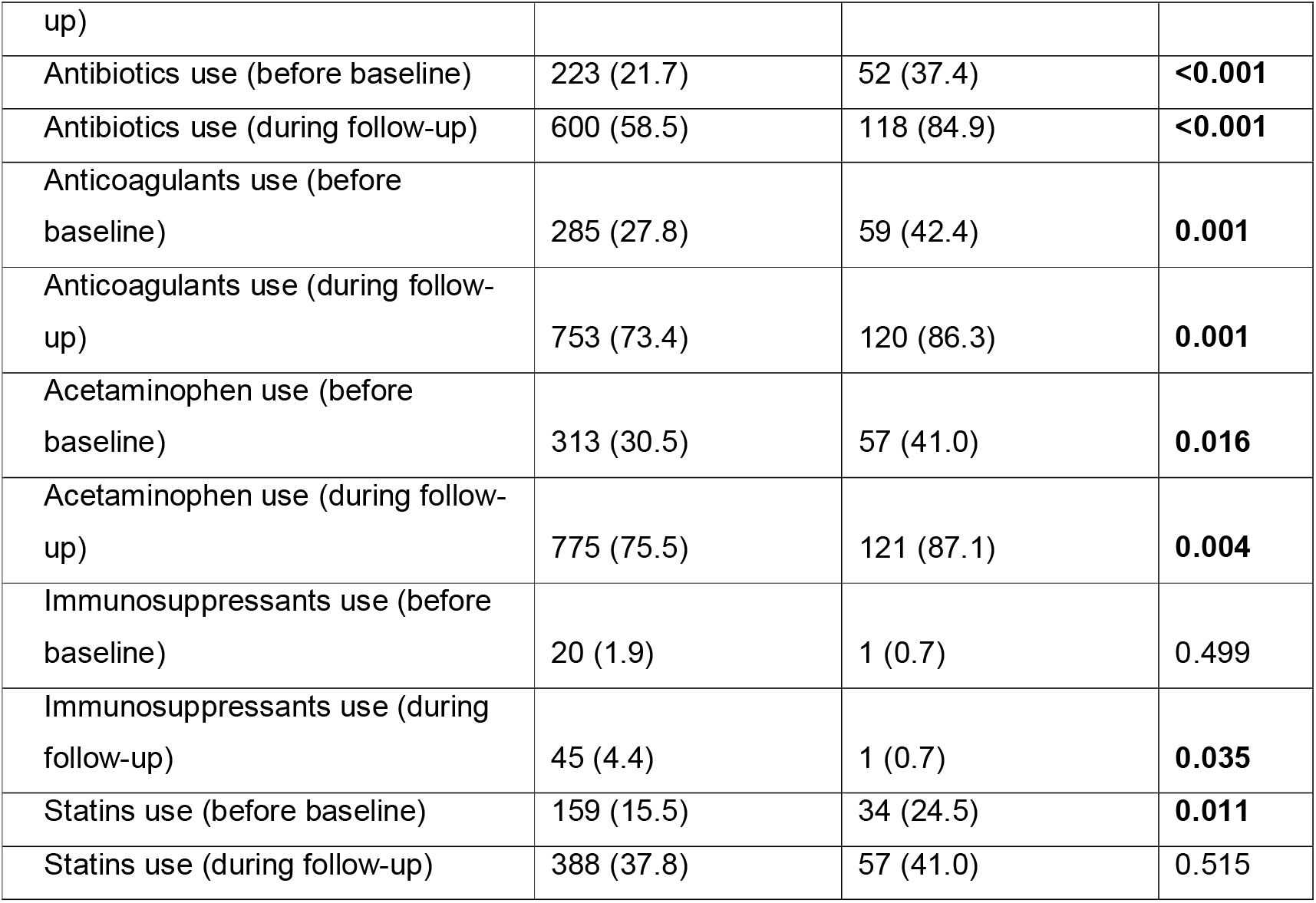
Patient characteristics, demographics, liver biochemistry, and clinical information in non-COVID-19 group, stratified by survival status. ^†^ 182 vs. 9 patients in survived subgroup vs. died subgroup had not all liver biochemistry baseline data available. * 179 vs. 9 patients in survived subgroup vs. died subgroup had not all liver biochemistry (excl. albumin) baseline data available at baseline. ^§^ 182 vs. 9 patients in survived subgroup vs. died subgroup had no data of ALT available at baseline. ^ 171 vs. 9 patients in survived subgroup vs. died subgroup had no data of ALP available at baseline. ^#^ 182 vs. 9 patients in survived subgroup vs. died subgroup had no data of bilirubin available at baseline. ^*^ 168 vs. 9 patients in survived subgroup vs. died subgroup had no data of albumin available at baseline. NB. For categorical variables, Fisher exact test was performed for comparison on cells with small counts (<5), otherwise Chi-square test was used. For continuous variables, Wilcoxon test was used for comparison due to non-normality. *ALT, Alanine transaminase; ALP, Alkaline phosphatase; CHD, Coronary heart disease; CKD, Chronic kidney disease; DM, Diabetes mellitus; HTN, Hypertension; IQR, Interquartile range; LLN, Lower limit of normal; ULN, Upper limit of normal*.

**Table S11.** Associations of baseline liver biochemistries with death among patients with COVID-19, investigated with Cox proportional-hazards modelling. ^†^ To make it more interpretable, we transfer the baseline albumin values to negative values. *NB*. Analyses were performed on the 492 COVID-19 patients with baseline liver biochemistry data available (survived vs. died: 346 vs. 146); variables (except for demographics, comorbidities) with p<0.1 in univariate analysis were included for multivariate analysis; in the multivariate analysis, HRs were fully adjusted for drugs use before baseline (including antiviral drugs, antibiotics, anticoagulants, acetaminophen, immunosuppressants, statins) to reduce confounding effects. *ALT, Alanine transaminase; ALP, Alkaline phosphatase; CHD, Coronary heart disease; CKD, Chronic kidney disease; DM, Diabetes mellitus; HTN, Hypertension; HR, Hazards ratio; IQR, Interquartile range*.

| Variables | Univariate analysis |  | Multivariate analysis |  |
| --- | --- | --- | --- | --- |
|  | Crude HR (95% CIs) | p-value | Adjusted HR (95% CIs) | p-value |
| Age (divided by 10 years) | 1.74 (1.53-1.97) | **<0.001** | **1.82 (1.56-2.13)** | **<0.001** |
| Gender (male) | 1.24 (0.91-1.71) | 0.18 | 1.25 (0.88-1.76) | 0.21 |
| Ethnicity (white) | 1.87 (1.25-2.8) | **0.002** | 1.09 (0.69-1.72) | 0.71 |
| Baseline ALT (IU/L) | 1 (0.99-1) | 0.39 |  |  |
| Baseline ALP (IU/L) | 1 (1-1) | 0.35 |  |  |
| Baseline bilirubin (umol/L) | 1.01 (1-1.02) | 0.18 |  |  |
| (- Baseline albumin) <sup>†</sup> (g/L) | 1.06 (1.04-1.09) | **<0.001** | **1.05 (1.02-1.09)** | **0.002** |
| Pre-existing Liver disease | 2.66 (1.44-4.9) | **0.002** | **3.15 (1.47-6.74)** | **0.003** |
| Pre-existing DM | 1.7 (1.16-2.48) | **0.006** | 1.43 (0.91-2.24) | 0.125 |
| Pre-existing HTN | 1.33 (0.96-1.84) | 0.085 | 0.66 (0.41-1.04) | 0.075 |
| Pre-existing CHD | 1.72 (1.11-2.65) | **0.015** | 1.02 (0.59-1.78) | 0.93 |
| Pre-existing CKD | 1.34 (0.83-2.17) | 0.23 | 0.76 (0.42-1.38) | 0.37 |
| Pre-existing Cancer | 1.71 (1.04-2.84) | **0.036** | 0.89 (0.48-1.65) | 0.71 |

**Table S12.** Associations of peak/nadir liver biochemistries with death among patients with COVID-19, investigated with Cox proportional-hazards modelling. ^†^ To make it more interpretable, we transfer the nadir albumin values to negative values. NB. Analyses were performed on the whole COVID-19 group excluding one died patient with missing data on death date; variables (except for demographics and comorbidities) with p<0.1 in univariate analysis were included for multivariate analysis; in the multivariate analysis, HRs were fully adjusted for drugs use before baseline (including antiviral drugs, antibiotics, anticoagulants, acetaminophen, immunosuppressants, statins) to reduce confounding effects. *ALT, Alanine transaminase; ALP, Alkaline phosphatase; CHD, Coronary heart disease; CKD, Chronic kidney disease; DM, Diabetes mellitus; HTN, Hypertension; HR, Hazards ratio; IQR, Interquartile range*.

| Variables | Univariate analysis |  | Multivariate analysis |  |
| --- | --- | --- | --- | --- |
|  | Crude HR (95% CIs) | p-value | Adjusted HR (95% CIs) | p-value |
| Age (divided by 10 years) | 1.74 (1.53-1.97) | **<0.001** | **1.81 (1.56-2.09)** | **<0.001** |
| Gender (male) | 1.24 (0.91-1.71) | 0.18 | 1.23 (0.88-1.73) | 0.23 |
| Ethnicity (white) | 1.87 (1.25-2.8) | **0.002** | 1.07 (0.7-1.64) | 0.75 |
| Peak ALT (IU/L) | 1 (1-1) | 0.35 |  |  |
| Peak ALP (IU/L) | 1 (1-1) | 0.73 |  |  |
| Peak bilirubin (umol/L) | 1.01 (1-1.01) | 0.21 |  |  |
| (-Nadir albumin) <sup>†</sup> (g/L) | 1.07 (1.05-1.09) | **<0.001** | **1.07 (1.04-1.10)** | **<0.001** |
| Pre-existing Liver disease | 2.66 (1.44-4.9) | **0.002** | **2.42 (1.18-4.95)** | **0.016** |
| Pre-existing DM | 1.7 (1.16-2.48) | **0.006** | 1.32 (0.84-2.09) | 0.23 |
| Pre-existing HTN | 1.33 (0.96-1.84) | 0.085 | 0.72 (0.47-1.11) | 0.13 |
| Pre-existing CHD | 1.72 (1.11-2.65) | **0.015** | 1.06 (0.63-1.79) | 0.82 |
| Pre-existing CKD | 1.34 (0.83-2.17) | 0.23 | 0.74 (0.42-1.3) | 0.29 |
| Pre-existing Cancer | 1.71 (1.04-2.84) | **0.036** | 1 (0.57-1.74) | 1 |

**Table S13.** Associations of baseline liver biochemistries with death among patients with COVID-19, investigated in subset/sensitivity analysis with Cox proportional-hazards models further adjusted for BMI. ^†^ To make it more interpretable, we transfer the baseline albumin values to negative values. *NB*. Analyses were performed on a subset of COVID-19 patients who had BMI and baseline liver biochemistry data (survived vs. died: 246 vs. 96); variables (except for demographics, comorbidities) with p<0.1 in univariate analysis were included for multivariate analysis; in the multivariate analysis, HRs were fully adjusted for drugs use before baseline (including antiviral drugs, antibiotics, anticoagulants, acetaminophen, immunosuppressants, statins) to reduce confounding effects. *ALT, Alanine transaminase; ALP, Alkaline phosphatase; CHD, Coronary heart disease; CKD, Chronic kidney disease; DM, Diabetes mellitus; HTN, Hypertension; HR, Hazards ratio; IQR, Interquartile range*.

| Variables | Univariate analysis |  | Multivariate analysis |  |
| --- | --- | --- | --- | --- |
|  | Crude | p-value | Adjusted | p-value |

**Table S13. Associations of baseline liver biochemistries with death among patients with COVID-19, investigated in subset/sensitivity analysis with Cox proportional-hazards models further adjusted for BMI.**
|  | HR (95% CIs) |  | HR (95% CIs) |  |
| --- | --- | --- | --- | --- |
| Age (divided by 10 years) | 1.71 (1.44-2.03) | **<0.001** | **1.79 (1.47-2.18)** | **<0.001** |
| Gender (male) | 0.95 (0.64-1.42) | 0.82 | 1.20 (0.79-1.83) | 0.39 |
| Ethnicity (white) | 1.92 (1.11-3.33) | **0.02** | 1.18 (0.66-2.09) | 0.58 |
| BMI | 0.98 (0.95-1.01) | 0.22 | 1.01 (0.97-1.04) | 0.72 |
| Baseline ALT (IU/L) | 1 (0.99-1) | 0.34 |  |  |
| Baseline ALP (IU/L) | 1 (1-1) | 0.41 |  |  |
| Baseline bilirubin (umol/L) | 1.01 (0.99-1.02) | 0.42 |  |  |
| (- Baseline albumin)<sup>†</sup> (g/L) | 1.06 (1.02-1.09) | **0.003** | **1.04 (1.01-1.09)** | **0.049** |
| Pre-existing liver disease | 3.15 (1.53-6.51) | **0.002** | **2.98 (1.25-7.14)** | **0.014** |
| Pre-existing DM | 1.86 (1.18-2.94) | **0.008** | 1.38 (0.80-2.37) | 0.25 |
| Pre-existing HTN | 1.44 (0.96-2.16) | 0.077 | 0.86 (0.48-1.52) | 0.60 |
| Pre-existing CHD | 1.33 (0.74-2.39) | 0.33 | 0.76 (0.37-1.53) | 0.43 |
| Pre-existing CKD | 1.30 (0.74-2.30) | 0.36 | 0.81 (0.40-1.62) | 0.54 |
| Pre-existing Cancer | 1.99 (1.09-3.65) | **0.026** | 1.05 (0.51-2.17) | 0.89 |

**Table S14.**
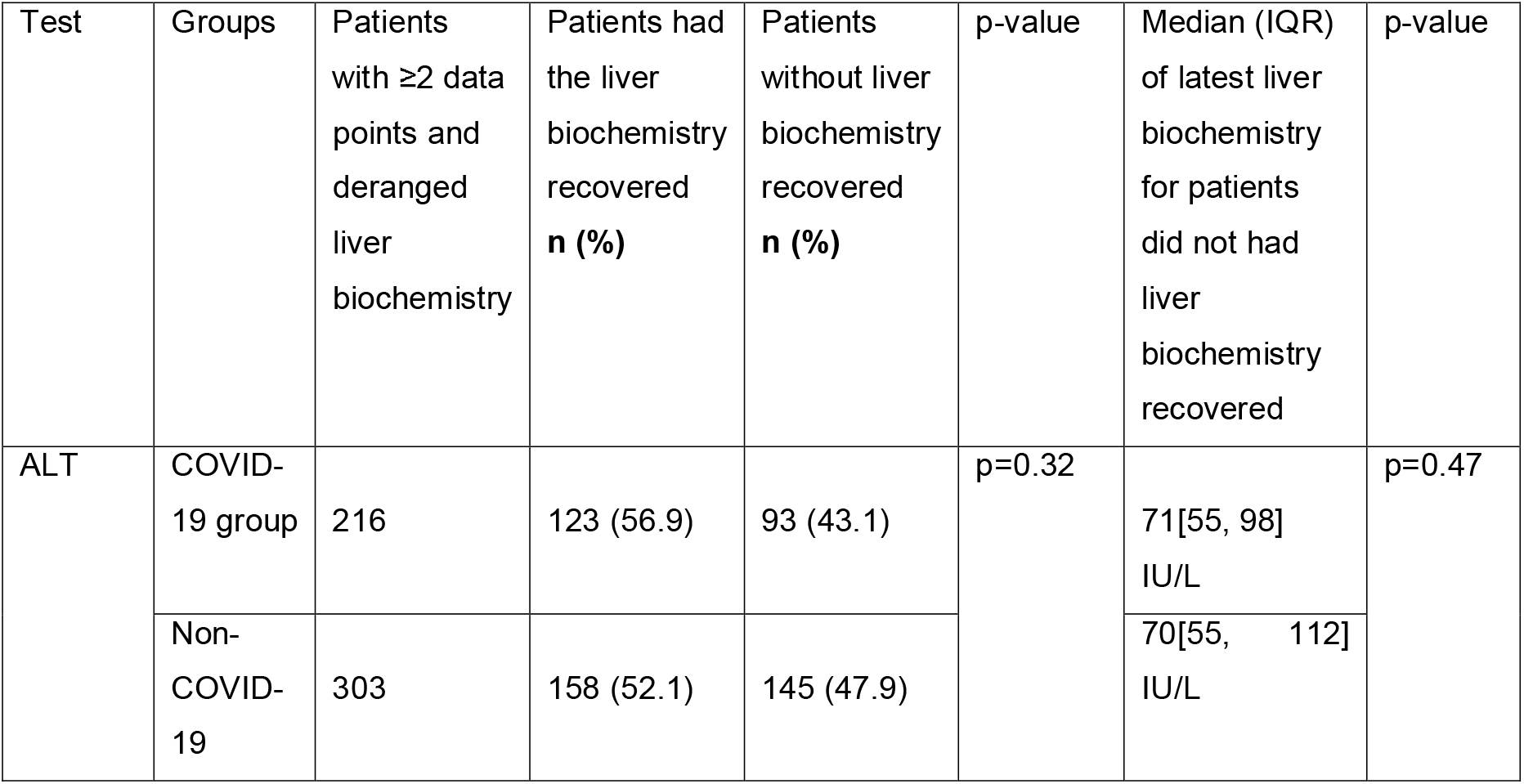
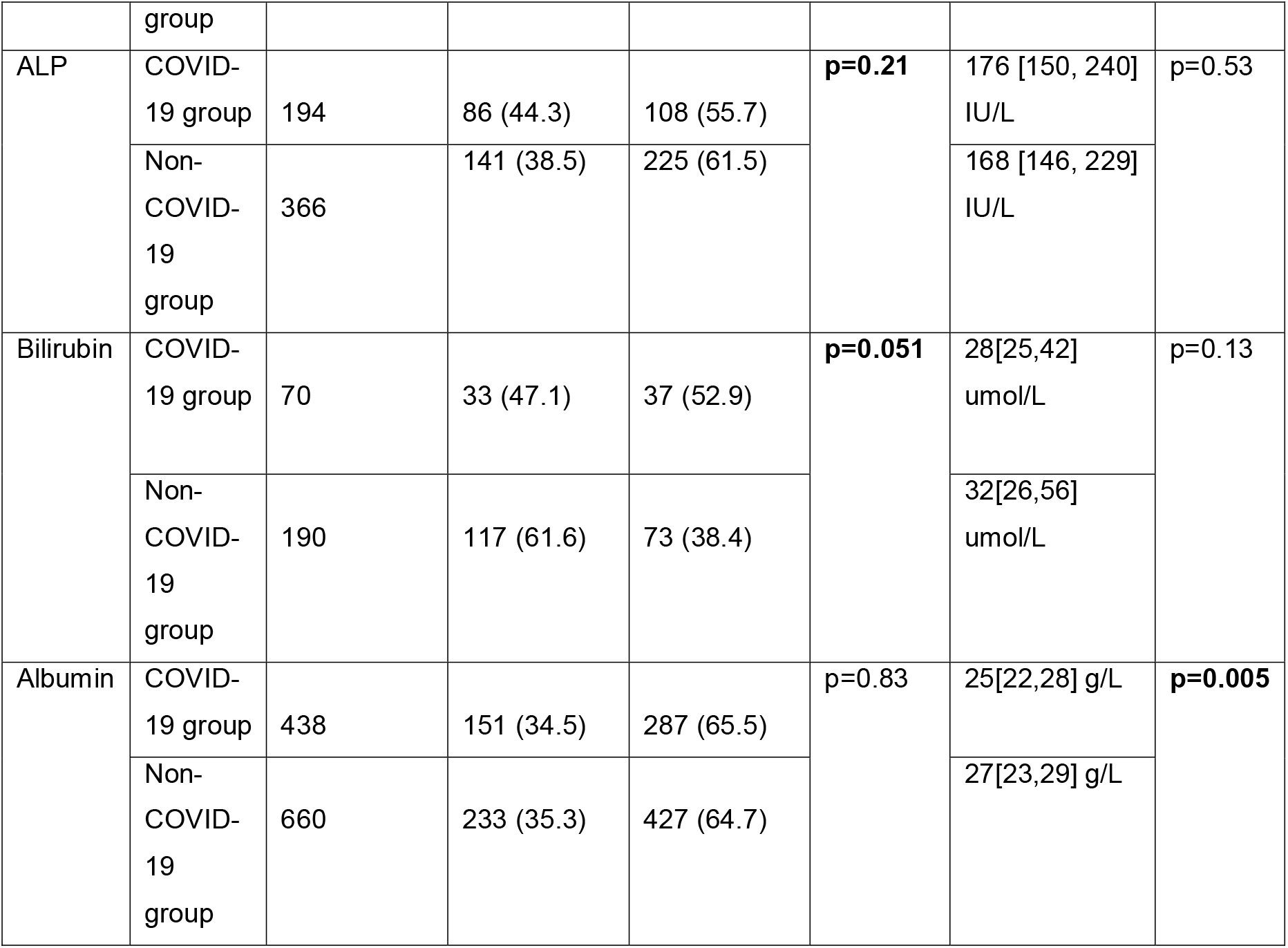
Detailed information regarding liver biochemistry recovery for each parameter in patients with and without COVID-19.

**Figure S1.**
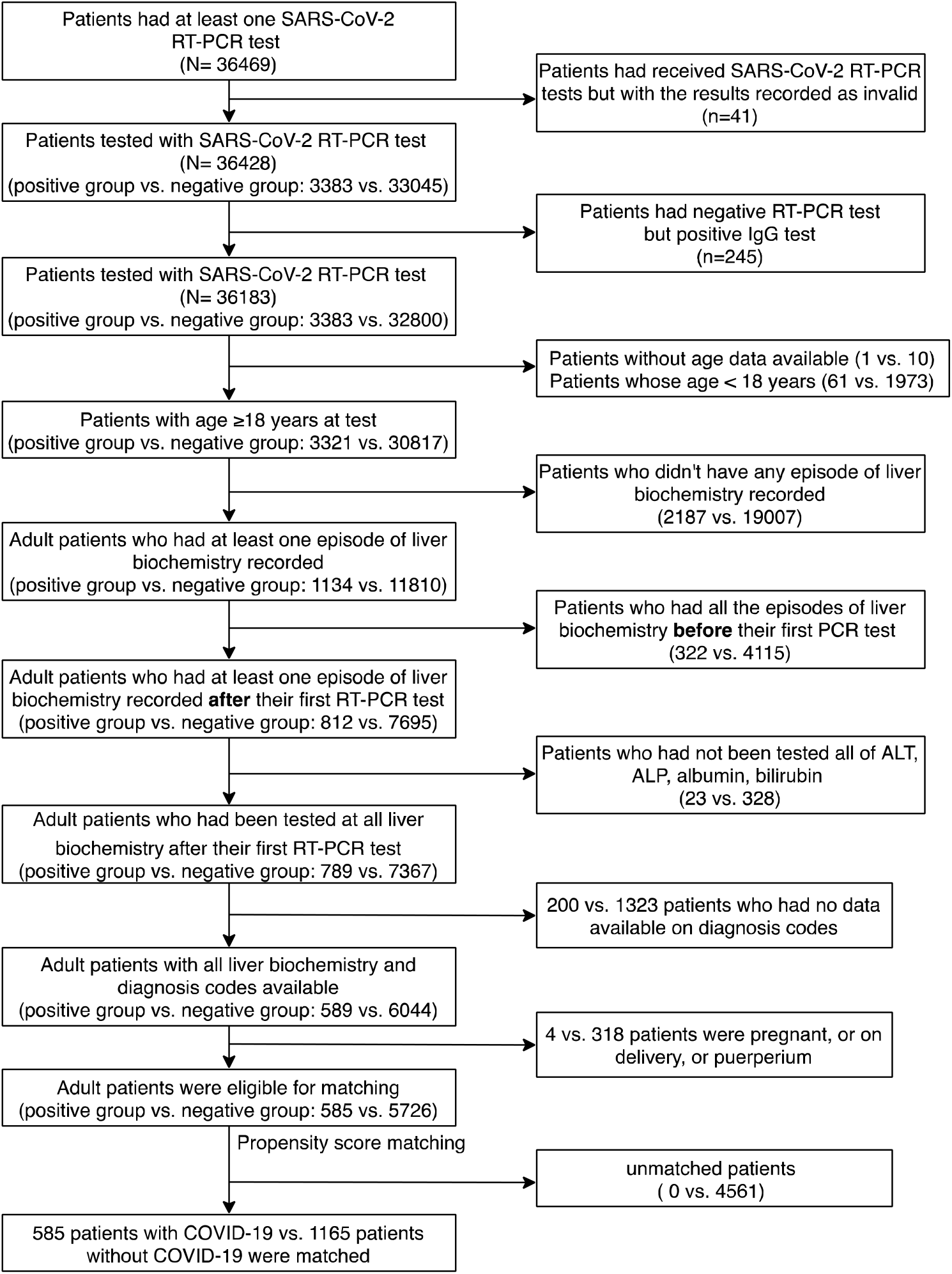

**Figure S2.**
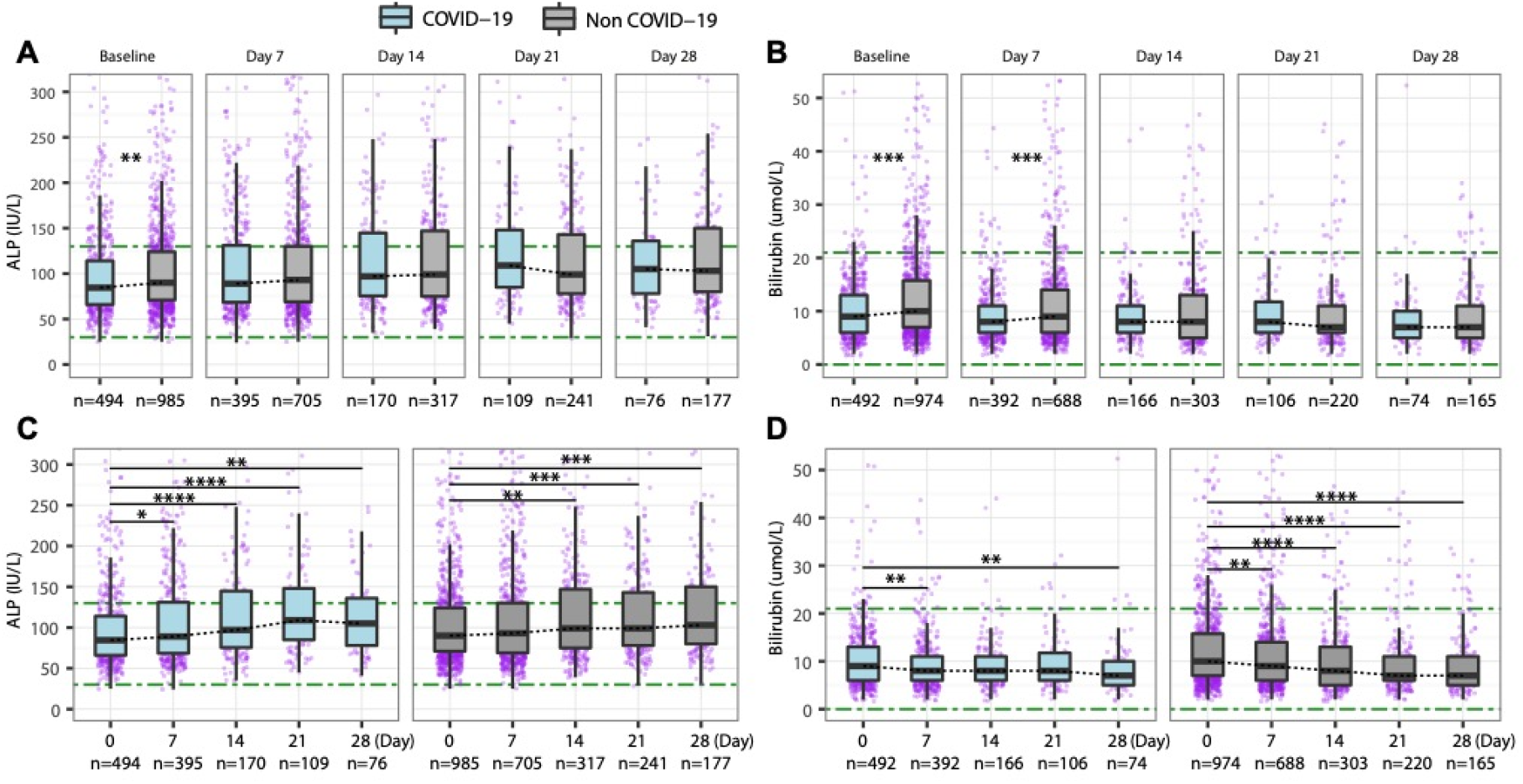
Comparison of liver biochemistry (ALP, bilirubin) between COVID-19 group and non-COVID-19 group at each time point and the longitudinal changes of liver biochemistry (ALP, bilirubin) over time within each group. (**A**) ALP comparison at baseline, 7, 14, 21, 28 days; (**B**) ALP changes over time; (**C**) Bilirubin comparison at baseline, 7, 14, 21, 28 days; (**D**) Bilirubin changes over time. *ALP, Alkaline phosphatase. Green dash-dotted lines indicate the lower limits of normal and the upper limits of normal. * p-value <0.05, ** p-value <0.01, *** p-value <0.001, **** p-value <0.0001*.

**Figure S3.**
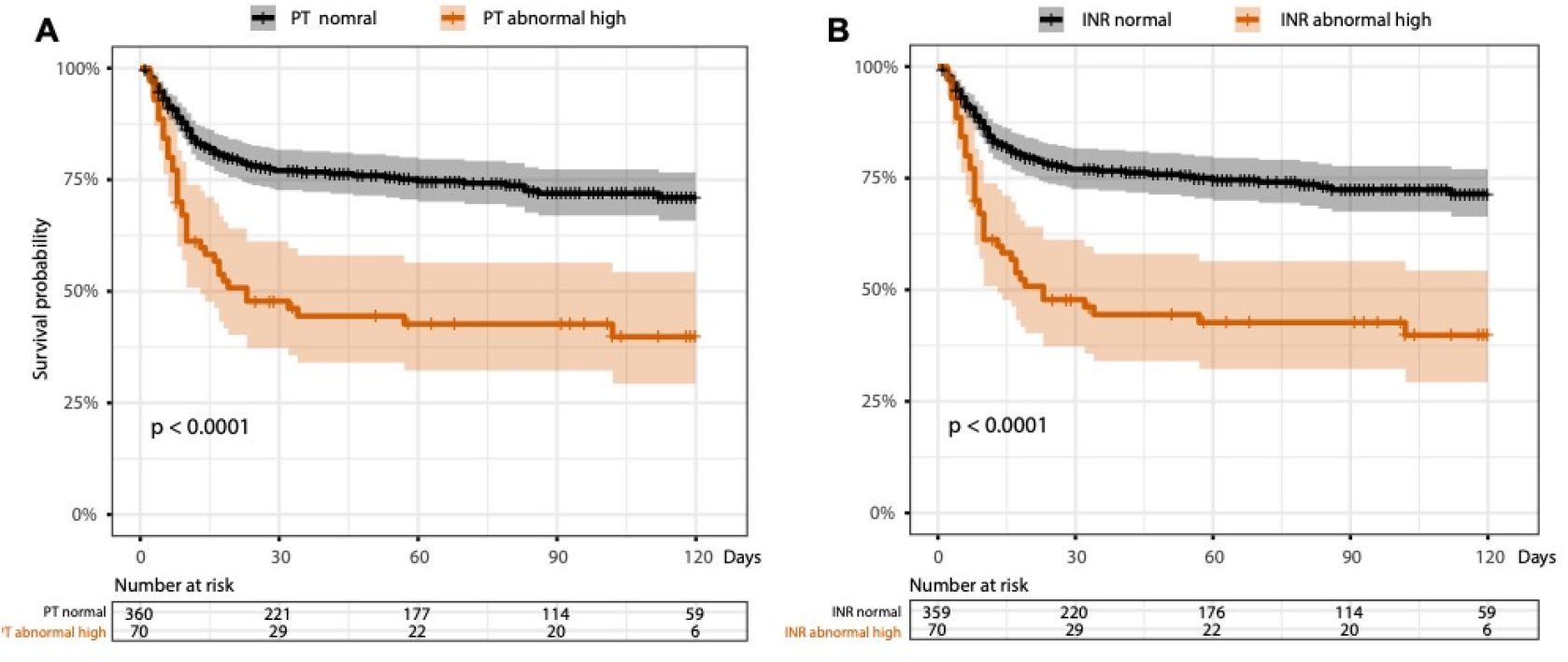
K-M curves for comparisonsof time from testing positivefor SARS-CoV-2 to death for subgroups stratified by. (**A**) Normal and abnormal high baseline PT; (**B**) normal and abnormal high baseline INR. *K-M, Kaplan-Meier; PT, Prothrombin time; INR, International normalised ratio. A subset of the COVID-19 group (n=430 vs. n=429) had data on PT and INR at baseline. p-values were based on the logrank test*.

**Figure S4.**
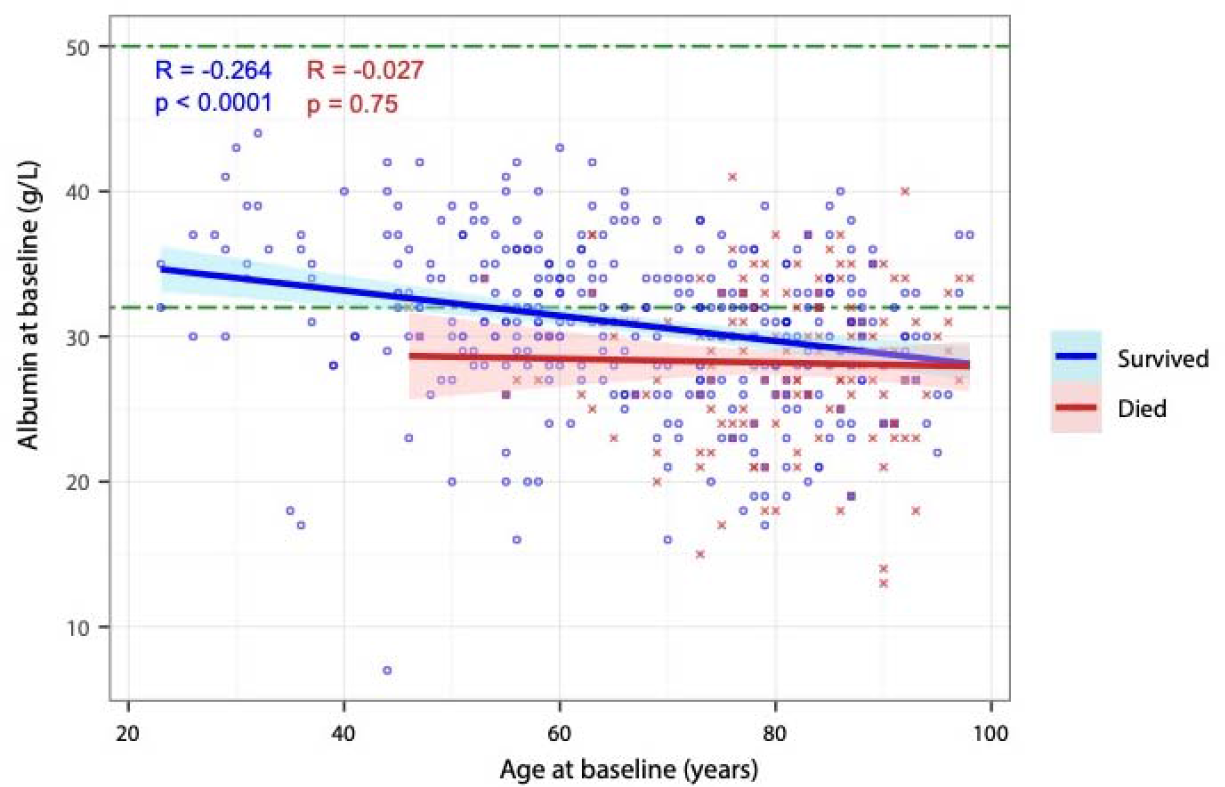
Correlation of age with baseline albumin at the time of RT-PCR test in the COVID-19 group, stratified by survival status at end of follow-up. *R represents Pearson’s correlation coefficient and p indicates linear regression significance*.

**Figure S5.**
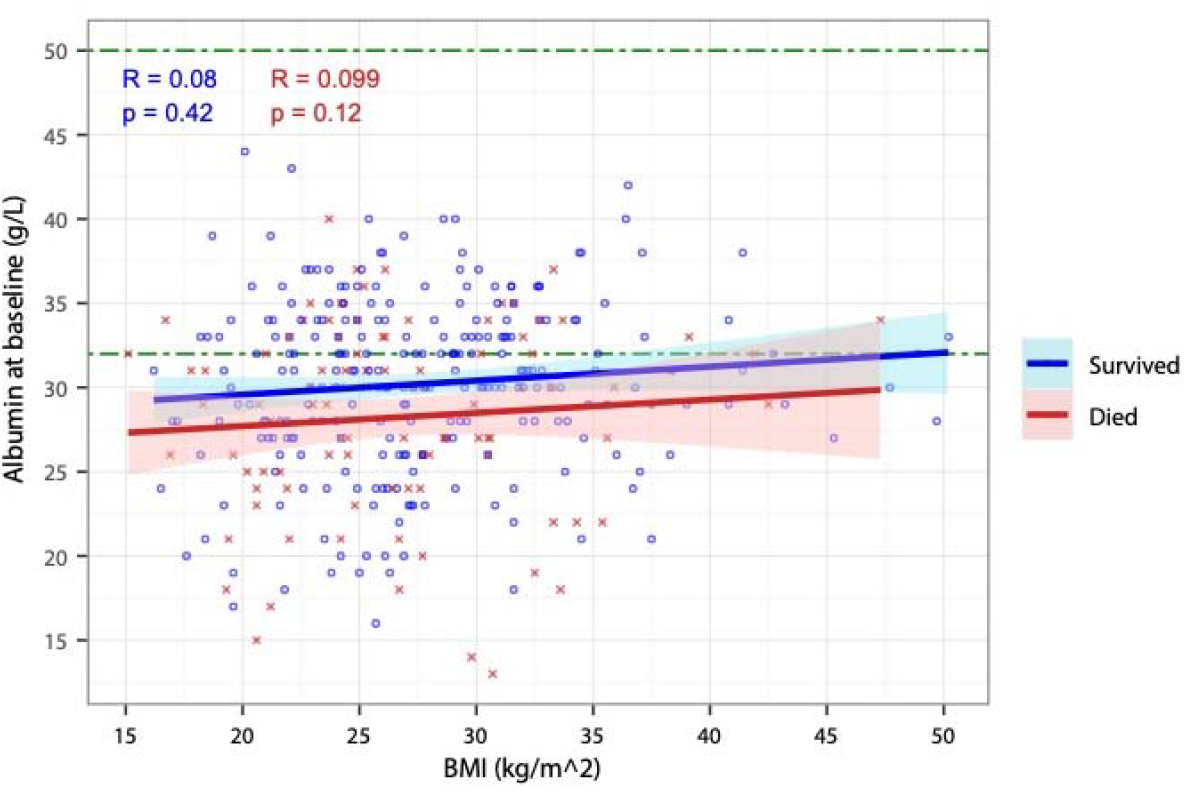
Correlation of body mass index (BMI) with baseline albumin at the time of RT-PCR test in the COVID-19 group, stratified by survival status at end of follow-up. *R represents Pearson’s correlation coefficient and p indicates linear regression significance*.

